# The Alzheimer’s Disease Diagnosis and Plasma Phospho-Tau217 (ADAPT) study stage 1: validating clinical cut-points against CSF and amyloid PET

**DOI:** 10.1101/2025.09.08.25335317

**Authors:** Ashvini Keshavan, Katharine Wiltshire, Ryan Wee, Irene Gorostiaga Belio, Katie Tucker, Melanie Hart, Michael P. Lunn, Michael C. B. David, Laura Rizzo, Omid Sadeghi-Alavijeh, Patricia Wilson, Daniel P. Gale, Amanda J. Heslegrave, Henrik Zetterberg, Nick C. Fox, Paresh Malhotra, Jonathan M. Schott

## Abstract

**INTRODUCTION:** We validated plasma p-tau217 cut-points for Alzheimer’s disease (AD) diagnosis using two commercial assays in two biomarker-defined cohorts and examined influences of pre-analytical factors and chronic kidney disease (CKD) on p-tau217 concentrations.

**METHODS:** Lumipulse (Fujirebio) and ALZpath (Quanterix) assays quantified plasma p-tau217 in symptomatic patients (AD status definition CSF n=257; amyloid PET n=76). ROC analyses established ≥95% sensitivity/specificity cut-points. In separate cohorts we evaluated the impact of pre-analytical handling/transport variations (n=40/10) and cognitively normal (CN)-CKD individuals (n=58).

**RESULTS:** Diagnostic accuracy was similar (AUROC Lumipulse 0.947; ALZpath 0.940). Lumipulse p-tau217 achieved 95% sensitivity and 97% specificity using dual cut-points (0.153/0.422 pg/mL), producing indeterminate results in 19.4% (CSF-defined) and 34.2% (PET-defined). P-tau217 concentrations were stable across handling conditions and kit lots, and mostly in the low-to-intermediate range in CN-CKD.

**DISCUSSION:** Lumipulse plasma p-tau217, now available in our UKAS-accredited clinical NHS laboratory, will be used in a randomized trial of p-tau217 result disclosure in memory services.

## 1. Background

Alzheimer’s disease (AD) diagnosis is typically formulated using clinical assessments and structural brain imaging, combined where available with molecular tools (for example measurement of CSF amyloid beta (Aβ) and phosphorylated tau (p-tau) or amyloid positron emission tomography (PET)). Introducing an effective blood test to support the diagnosis of AD would reduce the need for invasive CSF sampling and/or expensive amyloid PET and improve the accessibility of obtaining an early and accurate diagnosis in settings where those molecular tools are not currently available [1, 2].

There is strong evidence to show that plasma p-tau biomarkers are accurately able to discriminate AD neuropathology from other neurological diseases [3, 4]. In particular, plasma tau phosphorylated at threonine-217 (p-tau217), has been identified as a specific, high-performing biomarker for the differentiation of AD from other neurodegenerative diseases, when applied with appropriate high pre-test probabilities. It has higher diagnostic accuracy than other plasma biomarkers [5–7] and comparable performance against current CSF and PET clinical diagnostic standards [8–10]. Longitudinal studies have also shown that increases in plasma p-tau217 correlate with increased deficits in cognition and brain atrophy [11], and that elevated plasma p-tau217 is an accurate predictor of subsequent AD-related brain pathology, cognitive impairment and AD diagnosis [12–15]. Among commercially available plasma p-tau217 assays, the Lumipulse automated platform [5], and the ALZpath assay on the Single Molecule Array (Simoa) HD-X platform (Quanterix) [6, 16–19] have shown high performance for differentiating AD from non-AD. However, clinical cut-points need to be established and evaluated to ensure these assays can be reliably integrated into diagnostic pathways, minimizing the risk of misclassification and potential misdiagnosis.

The Alzheimer’s Disease Diagnosis and Plasma phospho-Tau217 (ADAPT) study is a UK multi-center study consisting of three stages. Stage 1 involved plasma p-tau217 assay selection and cut-point derivation. Stage 2 will examine the stability of those derived cut-points over time. Stage 3 will be a randomized controlled trial of the disclosure of plasma p-tau217 results to patients and clinicians, indexing proportion of AD diagnoses as the primary outcome.

In the first stage, described here, we compared the Lumipulse and ALZpath assays in a patient cohort against CSF AD biomarker status, and determined the optimum cut-points to apply clinically using a dual cut-point approach [20]. We then validated these p-tau217 assay cut-points against amyloid PET status in an independent cohort. Secondary aims within stage 1 were to consider the impact of comorbidities [21] by identifying any associations of chronic kidney disease (CKD) and body mass index (BMI) with p-tau217 measurements taken from participants within this study and from a separate cohort of participants with stage 3-4 CKD, and to investigate the effects of varied pre-analytical sample handling factors [22] and assay kit lot variability on p-tau217 measurements, so as to guide sample handling protocols for the stage 3 trial.

## 2. Methods

### 2.1 CSF cohort

#### 2.1.1 Study participants and clinical classification

Participants were identified from an existing cohort of patients who attended the cognitive clinic at University College London Hospitals NHS Foundation Trust, National Hospital for Neurology and Neurosurgery (London, UK), between August 2017 and September 2024, and were referred for lumbar punctures to investigate their cognitive symptoms. Informed consent, or assent in discussion with participants’ consultees, was obtained for all participants to participate in biomarker research studies and for researchers to have access to medical records. Ethical approval for use of the samples in ADAPT stage 1 was provided by an existing study (Wolfson CSF study 12/0344, NRES London Queen Square, August 2013, PI Schott). Participants were included if minimum stored volumes of 1mL of EDTA plasma and 500µL of matched CSF were available.

Participants were classified as AD biomarker status positive if CSF Aβ_42_/Aβ_40_ ratio was ≤0.065 (“amyloid only” definition), measured using the Lumipulse G1200 immunoassay platform (Fujirebio) in clinical routine. Sensitivity analyses were undertaken using two further definitions of AD status. The “amyloid and p-tau” definition was CSF Aβ_42_/Aβ_40_ ratio ≤0.065 AND p-tau181 ≥57pg/mL. The “clinical AD status” was based on the most recent clinical diagnosis confirmed via clinical follow-up and informed by results of CSF testing using whichever AD biomarker assay combination was in clinical use at the time of testing, as between August 2017 and March 2020 the Innotest (Fujirebio) ELISA for Aβ42 and total tau were used, and between April 2020 and September 2024 the aforementioned Lumipulse CSF assays for Aβ_42_/Aβ_40_ and p-tau181 were used.

To assess the impact of comorbidities, information on kidney function (serum creatinine, estimated glomerular filtration rate (eGFR, calculated using the CKD-EPI 2021 equation [23]), and resulting CKD stage) within ±6 months and BMI within ±1 year of the lumbar puncture were collected from medical records where available and used in this study. Basic demographic data were also recorded including age at sampling, sex and ethnicity (where specified in the medical record).

#### 2.1.2 Blood sample collection and processing

Blood was collected by peripheral venipuncture from each participant into 8mL K2-EDTA tubes and CSF was collected by lumbar puncture using the protocol previously reported [24]. Samples were processed within 2 hours of collection. Blood was centrifuged at 2000 *g* for 10 minutes at room temperature and CSF was centrifuged at 1750 *g* for 5 mins at 4°C. Plasma and CSF were aliquoted and stored at −80°C until analysis.

### 2.2 Amyloid PET cohort

#### 2.2.1 Study participants and clinical classification

Participants were identified from an existing cohort of patients who attended the cognitive clinic at Imperial College Healthcare NHS trust, Charing Cross Hospital (London, UK), between August 2020 and November 2024 for an amyloid PET brain scan as part of other standard diagnostic procedures (the Imperial Amyloid PET Cohort Study 20/LO/0442, NRES London Camden and Kings Cross UK Research Ethics Committee, June 2020, PI Malhotra). An additional set of participants from the same ongoing cohort who attended the clinic between April and July 2025 were included in a sample transport temperature comparison study, described in section 2.2.2 below. Informed consent to participate in biomarker research studies was obtained from all participants. Participants were included if a minimum of 1mL of stored EDTA plasma was available.

Visual read amyloid burden was reported by two nuclear medicine experts and classified as amyloid positive or negative as previously reported, with amyloid load scores assigned as: 1 = no amyloid load, 2 = mild amyloid load, and 3 = significant amyloid load [25, 26].

To assess the impact of comorbidities, information on kidney function (serum creatinine, eGFR, and resulting CKD stage) within ±12 months of the date of the amyloid PET scan was collected from medical records where available and used in this study. BMI data were not available from this cohort. Basic demographic data were also recorded including age at PET scan, sex and ethnicity (where specified in the medical record).

#### 2.2.2 Blood sample collection and processing

Blood was collected by peripheral venipuncture from each participant into 4mL K3-EDTA tubes. Samples were processed within 2 hours of extraction. Blood was centrifuged at 2000 *g* for 10 minutes at 4°C. Plasma was aliquoted and stored at −80°C until analysis.

For the transport temperature comparison samples, immediately after plasma centrifugation, plasma samples were placed in intermediate storage either at −20°C for a maximum of two weeks and transported in a Bio-Freeze temperature controlled transport bottle system (Bio-packaging Ltd, Coventry, UK) for <24 hours at −20-0°C to UCL, or at −80°C for a maximum of two weeks and transported for <24 hours on dry ice to UCL. Samples were then stored at −80°C on arrival at UCL prior to analysis.

### 2.3 Chronic kidney disease cohort

#### 2.3.1 Study participants and clinical classification

Participants were identified from an existing cohort of patients with chronic kidney disease (CKD) due to autosomal dominant polycystic kidney disease (ADPKD) at the Royal Free London NHS Foundation Trust, Royal Free Hospital (London, UK), who provided plasma samples between 2012-2018. Use of plasma samples for this study was approved under ethics for the Polycystic Kidney Disease (PKD) Biobank, sponsored by PKD Charity at Royal Free (05/Q0508/6). Informed consent to participate in research was obtained from all participants. CKD stages were classed according to serum creatinine-based estimated glomerular filtration rate (eGFR, in ml/min/1.73m^2^ calculated using the CKD-EPI 2021 equation [23]): G1 = eGFR ≥ 90 (normal), G2 = eGFR 60-89 (mild decrease), G3a = eGFR 45-59 (mild to moderate decrease), G3b = eGFR 30-44 (moderate to severe decrease), G4 = eGFR 15-29 (severe decrease), G5 = eGFR <15 (kidney failure). Samples were included in this study if participants were aged <60 years at blood sampling (as this would provide an expected background prevalence of cerebral amyloid deposition in these cognitively normal individuals of <20% [27]), CKD stage was G1-G4 and at least 250uL of stored EDTA plasma was available. Basic demographic data were also recorded for all participants including age at sampling, sex, BMI and ethnicity (where available from medical records).

#### 2.3.2 Blood sample collection and processing

Blood was collected by peripheral venipuncture from each participant into K2-EDTA tubes. Samples were processed within 2 hours of venesection. Blood was centrifuged at 1000 *g* for 10 minutes at room temperature. Plasma was aliquoted and stored at −80°C until analysis.

### 2.4 Pre-analytical sample handling experiments

#### 2.4.1 Study participants

Participants were recruited from the cognitive neurology clinic at the National Hospital for Neurology and Neurosurgery (London, UK), under the provisions of two existing studies – the Wolfson CSF study (12/0344, NRES London Queen Square, August 2013, PI Schott; patients only) and the Biomarkers and Genetics in Dementia study (03/N049, NRES London April 2003, PI Fox; patients and healthy volunteers). All participants provided written informed consent. Basic demographic data were recorded including age at sampling and sex.

#### 2.4.2 Sample collection and processing

Blood samples were collected from participants by peripheral venipuncture into 8 mL K2-EDTA tubes and handled according to pre-specified conditions that varied pre- and post-centrifugation delays, storage temperatures and number of freeze-thaw cycles. For all sets of experiments, blood samples were centrifuged at 2000 *g* for 10 minutes at room temperature and plasma was stored at −80°C until analysis. A total of 40 participants provided blood samples, to allow for 10 per experimental condition. Plasma from two participants were excluded from the final analysis due to incorrect sample processing at the centrifugation stage. Four experiments were conducted to study the following pre-analytic handling factors:

*Experiment 1 - pre-centrifugation delay at room temperature:* Following receipt of blood samples in the laboratory, blood samples were left to stand in their original collecting EDTA tubes at room temperature for either 30 minutes (baseline condition) versus 1 hour or 3 hours from the time of venipuncture before centrifugation. The supernatant plasma was then aliquoted and stored immediately in −80°C.

*Experiment 2 - pre-centrifugation delay at 2-8°C:* Blood samples were left to stand in their original collecting EDTA tubes for either 30 minutes at room temperature (baseline condition) versus 3 hours or 24 hours in a refrigerator at 2-8°C from the time of venipuncture prior to centrifugation. The supernatant plasma was then aliquoted and stored immediately in −80°C.

*Experiment 3 - post-centrifugation delay durations and storage temperatures:* Blood samples stood in EDTA tubes at room temperature for 30 minutes from the time of venipuncture, then underwent centrifugation and aliquoting of the supernatant plasma into polypropylene tubes. Aliquots were left to stand under the following conditions that varied post-centrifugation delay durations under different temperatures: no delay (baseline condition) versus 4 hours room temperature, 24 hours room temperature, 4 hours at 2-8°C, 24 hours at 2-8°C, 2 weeks at 2-8°C, or 2 weeks at −20°C. After the assigned delay durations had elapsed, the samples were stored in −80°C.

*Experiment 4 - freeze-thaw cycles:* Blood samples stood in EDTA tubes for 30 minutes from the time of venipuncture at room temperature before undergoing centrifugation, aliquoting of the supernatant plasma and storage in −80°C. Following storage at −80°C for at least 24 hours, aliquots were retrieved from storage and left to stand in room temperature to thaw for at least 1 hour before being replaced back into −80°C. These freeze-thaw cycles were repeated to generate the following number of freeze-thaw cycles: 1 (baseline condition) versus 2, 3, and 4 cycles. Samples were stored in −80°C for at least 24 hours in between freeze-thaw cycles.

### 2.5 Plasma p-tau217 assays

#### 2.5.1 Lumipulse

Plasma samples were tested blinded in singlicate on the Lumipulse G1200 fully automated immunoassay platform using the commercially available Lumipulse G plasma p-tau217 immunoreaction cartridge kits (Fujirebio Europe, Ghent, Belgium) according to manufacturer instructions. Prior to analysis, samples were thawed at room temperature, vortexed briefly and centrifuged at 2000 *g* for 10 minutes at room temperature, and the supernatant pipetted into Hitachi analyzer sample cups for loading into the Lumipulse platform. All samples were quantifiable above the lower limit of quantification (LLOQ) of 0.03pg/mL provided by the manufacturer, which had also been independently verified by the laboratory using double dilution of a low p-tau217 plasma sample to this concentration, with serial quantification remaining within a CV of <10%.

#### 2.5.2 ALZpath

Plasma samples were tested blinded in singlicate (pre-analytic experiments) or duplicates (main CSF and amyloid PET cohort samples) on the Single Molecule Array (Simoa) HD-X immunoassay platform using the ALZpath p-tau217 v2 assay kit according to manufacturer instructions (Quanterix, MA, US). Prior to analysis, samples were thawed at room temperature, vortexed briefly, and centrifuged at 10 000 *g* for 5 minutes at room temperature prior to plating and loading into the HD-X platform. All samples were quantifiable above the LLOQ of 0.06pg/mL provided by the manufacturer.

#### 2.5.3 Lumipulse lot-to-lot variability experiments

A subset of samples from the CSF cohort (detailed in section 2.1) were selected for a lot-to-lot variability experiment using two lots of version 1 (v1) and two lots of version 2 (v2) Lumipulse G plasma p-tau217 kits (Fujirebio). Plasma samples were tested on the Lumipulse G1200 platform using three p-tau217 kit lots. Samples were thawed, briefly vortexed and centrifuged at 2000 *g* for 10 mins prior to testing on cartridge lot #5066 (v1), #5084 (v2) and #5086 (v2). When comparing the data for each cartridge lot, additional data from the same subset of plasma samples, tested with kit lot #4128 (v1) for the CSF cohort (described in section 2.5.1), were included in the analysis for lot-to-lot variability.

### 2.6 Statistical analysis

#### 2.6.1 Descriptive statistics

All statistics and plots were calculated and created using custom scripts written in R studio v2024.12.0 (Posit Software, PBC), excepting the pre-analytical sample handling data (details in section 2.6.5 below). All plasma p-tau217, serum creatinine and BMI data were tested for normality using a Shapiro-Wilk test, and based on these tests, data were assumed to be not normally distributed, indicating the use of non-parametric further statistical testing.

To report participant demographics, continuous variables were summarized using mean and standard deviation (SD) (participant age), or median and interquartile range (IQR) (Lumipulse/ALZpath plasma p-tau217 concentrations (pg/mL), serum creatinine, eGFR, BMI). Categorical and binary variables (sex, AD status, ethnicity, CKD stage) were summarized using percentages.

#### 2.6.2 CSF cohort

Spearman’s rank correlation coefficient *rho* (R) was used to describe the correlation of the Lumipulse and ALZpath p-tau217 assay measurements. For each assay, median fold-change in p-tau217 measurements between non-AD and AD status participants were calculated and tested for statistically significant differences using a Wilcoxon signed-rank test. For assessment of diagnostic performance of the two assays, receiver operating characteristic (ROC) analysis was performed on plasma p-tau217 measurements using AD status (non-AD or AD), classified using the CSF “amyloid only” definition (described in section 2.1.1) to calculate the area under the curve (AUC) with 95% CI for both the Lumipulse and ALZpath assays after formulating logistic regression models including either p-tau217 assay as a sole independent variable, or additionally including covariates of age, sex, serum creatinine and BMI. AUCs for each model with addition of variables were tested for statistically significant differences in comparison with the base model including age and sex alone using DeLong tests, and Akaike Information Criteria (AIC) were used to ascertain whether addition of covariates made significant differences to model prediction, with AIC reduction of 20 or more deemed as significant. Cut-points were derived from the ROC analyses using p-tau217 alone, using a dual cut-point approach [5, 16, 20] for 95% sensitivity and 95% specificity. The percentage of individuals with plasma p-tau217 results in the intermediate zone was evaluated, and confusion matrices were used to identify the number of false positive and false negative results, allowing for calculation of negative (NPV) and positive predictive values (PPV) with 95% CI and overall test accuracy for those individuals assigned a low or high value. We also evaluated a cut-point combination that maintained 95% sensitivity but maximized specificity while keeping the percentage of individuals in the intermediate zone at <20%. Cut-points for 90% sensitivity and 90% specificity, and 97.5% sensitivity and 97.5% specificity were also calculated for comparison of cut-point metrics.

Sensitivity analyses were undertaken whereby the derived 95% sensitivity and maximized specificity cut-points were also assessed for their NPV, PPV and accuracy using two other definitions of AD status – the CSF “amyloid and p-tau” definition and the “clinical AD status” (detailed in section 2.1.1).

#### 2.6.3 Amyloid PET cohort

Correlation of the Lumipulse and ALZpath assay p-tau217 measurements, fold changes and ROC analyses against amyloid PET visual read status were undertaken similarly to the statistical procedures described for the CSF cohort in section 2.6.2.

The 95% sensitivity and maximized specificity cut-points calculated from the CSF cohort (classified using CSF Aβ_42_/Aβ_40_ ratio only) were then applied to the amyloid PET cohort. The percentage of individuals with plasma p-tau217 results in the intermediate zone was evaluated, confusion matrices were used to identify the number of false positive and false negative results and calculate the negative (NPV) and positive predictive values (PPV) with 95% CI and overall accuracy.

A Passing-Bablok regression with Pearson’s correlation coefficient (*r*) was used to analyze differences in plasma p-tau217 following post-centrifugation storage and transport at −20°C versus −80°C.

#### 2.6.4 CKD cohort

Lumipulse plasma p-tau217 data for each CKD stage 1-4 was plotted against AD and non-AD plasma p-tau217 values and statistical significance of the difference in values between the AD group and the non-AD group and the non-AD group and each CKD group was tested using a Wilcoxon signed-rank test. Pooling together the samples from the CSF and CKD cohorts, multiple linear regression models were used to assess associations of serum creatinine or CKD stage and BMI with log-transformed p-tau217, adjusted for age, sex and cohort.

#### 2.6.5 Pre-analytical handling experiments

P-tau217 data were expressed as a relative (percentage) change compared to the base condition in each experiment. Friedman tests were used as a non-parametric one-way repeated-measures method to assess the null hypothesis that there was no between-group difference in the distribution of relative p-tau217 levels across pre-analytical handling conditions. Analyses were conducted in Python 3.6 on the Jupyter Notebook platform using the *scipy* and *scikit_posthocs* modules for statistical analysis and custom-written code. Where the null hypothesis was rejected at α = 0.05, we proceeded with a Nemenyi post-hoc test to identify the experimental groups contributing to the between-group differences. Further, we chose a ±10% change from baseline condition as a likely clinically significant change in relative p-tau217 levels, similar to previous publications [28–30]. Individual-level CVs in the relative changes of plasma p-tau217 compared to the base condition were calculated as the standard deviation of each subject’s relative p-tau217 fluctuations (excluding the base condition) divided by the mean (excluding the base condition); individual-level CVs were plotted against absolute base p-tau217 levels to examine the relationship between p-tau217 levels and the relative variability of p-tau217 measurements across pre-analytical handling conditions. Data presented as median ±IQR, unless otherwise stated.

#### 2.6.6 Lumipulse lot-to-lot variability experiments

Spearman’s rank correlation coefficient *rho* (R) was used to describe the correlation of the Lumipulse p-tau217 assay measurements between the four kit lots, with p-values for statistical significance of the correlation.

## 1. Results

### 3.1 Participant demographic characteristics

#### 3.1.1 CSF cohort

The CSF cohort included 257 individuals with a mean age at sampling of 63.3 years (SD 7.3 years); 153 (59.5%) were male; 159 (61.9%) were AD status positive, defined as CSF Lumipulse Aβ_42_/Aβ_40_ ratio ≤0.065 only; 112 (43.6%) identified their ethnicity as White, 18 (7.0%) as Black, 5 (2.0%) as Asian, 5 (2.0%) as Other, and 117 (45.5%) did not have their ethnicity specified in medical records. Serum creatinine was available from the medical records within ±6 months of plasma sampling from 62 participants (24.1%), in whom median serum creatinine was 74 (IQR 63-87) µmol/L and CKD stages are detailed in Table 1. BMI data were available from 151 participants (58.8%), in whom median BMI was 25.8 (IQR 22.8-29.1). Most recent clinical diagnosis data was available for all participants and was categorized into AD and other non-AD categories detailed in Table 1.

**Table 1:**
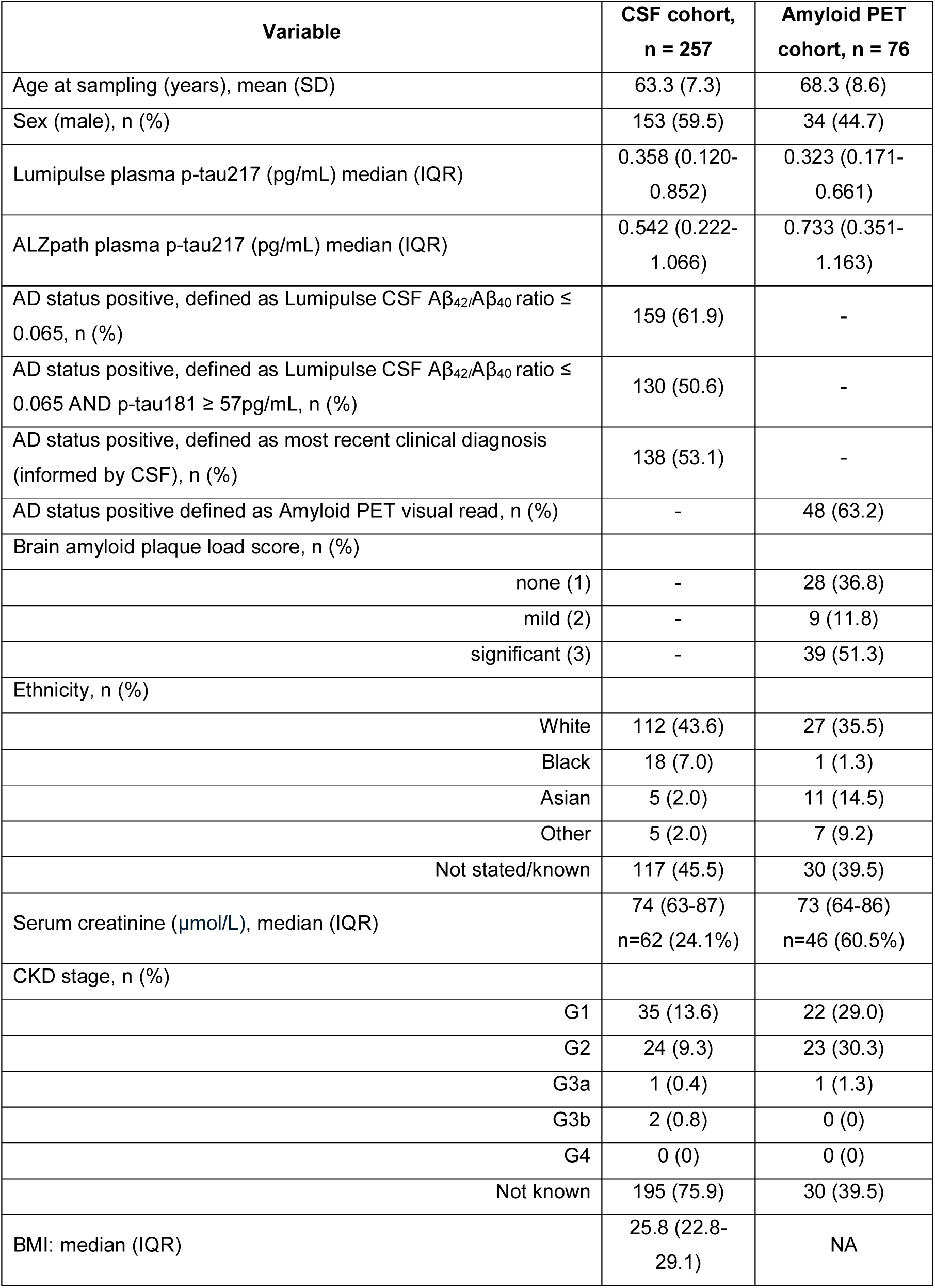
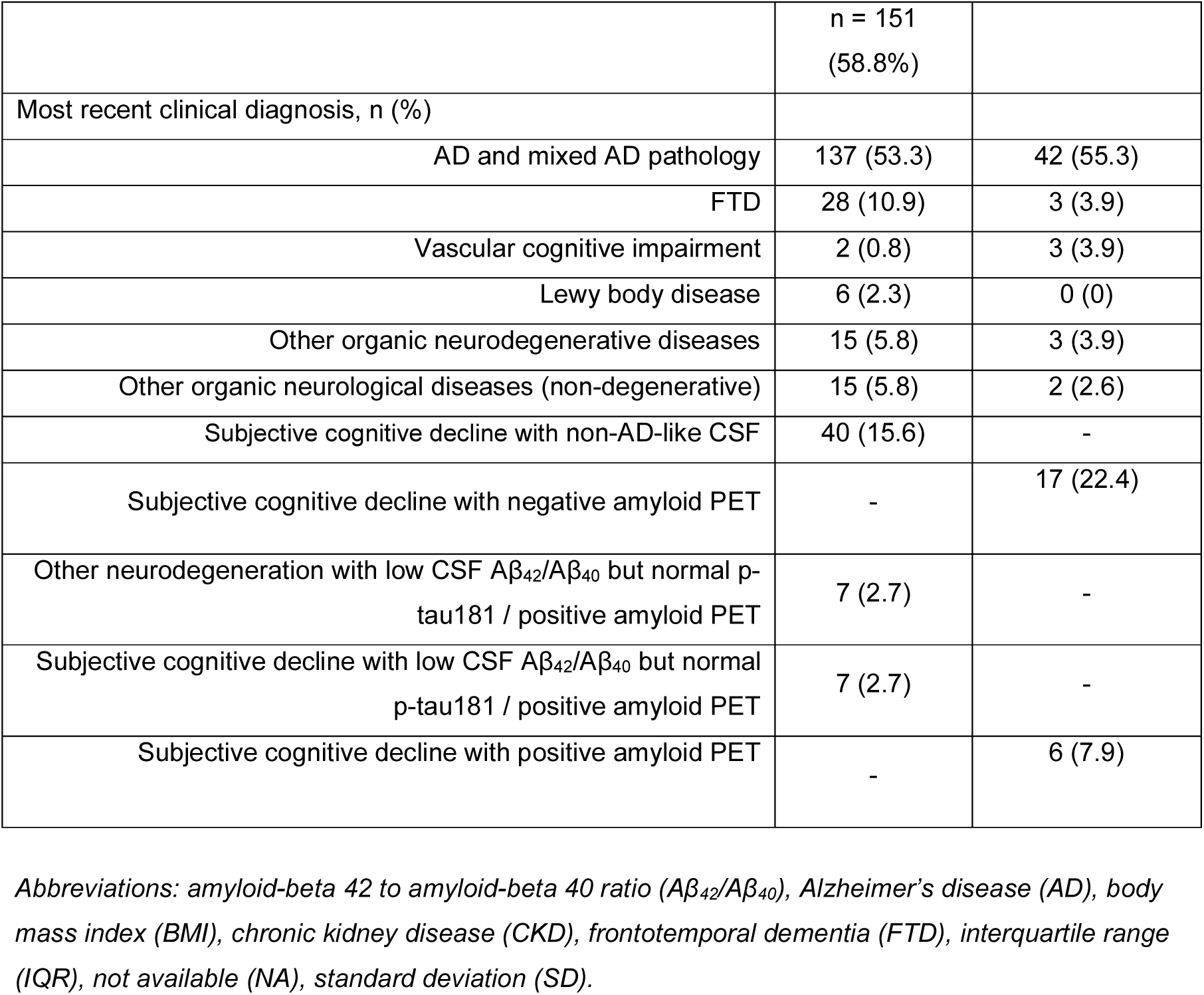
Participant demographic characteristics for the CSF and amyloid PET cohorts.

#### 3.1.2 Amyloid PET cohort

The amyloid PET cohort included 76 individuals with a mean age at PET scan of 68.3 years (SD 8.6 years); 34 (44.7%) were male; 48 (63.2%) were amyloid PET visual read positive, with brain amyloid plaque load scores of none (1) in 28 (36.8%), mild (2) in 9 (11.8%) and significant (3) in 39 (51.3%) participants. Ethnicity was identified as White in 27 individuals (35.5%), Black 1 (1.3%), Asian 11 (14.5%) Other 7 (9.2%) and 30 individuals (39.5%) did not have their ethnicity specified in medical records. Serum creatinine was available from the medical records within ±1 year of plasma sampling in 46 participants (60.5%) in whom median serum creatinine was 73 (64-86) µmol/L and CKD stages are detailed in Table 1. Most recent clinical diagnosis data was available for all participants and was categorized into AD and other non-AD categories detailed in Table 1.

#### 3.1.3 CKD cohort

The CKD cohort included 58 individuals below the age of 60 years, with a mean age at sampling of 44.4 years (SD 8.5 years); 32 (55.2%) were male; 27 (46.5%) identified their ethnicity as White, 3 (5.2%) as Black, 3 (5.2%) as Asian, 6 (10.4%) as Other, and 19 (32.8%) did not have their ethnicity specified in medical records. Numbers of participants in each CKD stage 1-4 were CKD-1: 11 (19.0%), CKD-2: 15 (25.9%), CKD-3a: 16 (27.6%), CKD-3b: 9 (15.5%), and CKD-4: 7 (12.1%). BMI data were available from 40 participants (69.0%), in whom median BMI was 25.0 (IQR 24.0-28.8) (Supplementary Table 1).

### 3.2 Plasma p-tau217 assay performance comparison

#### 3.2.1 CSF cohort

Lumipulse and ALZpath p-tau217 assay measurements were highly correlated (*rho* 0.86, p<0.001) (Supplementary Figure 1a).

Using the Lumipulse assay AD classification using the CSF “amyloid only” definition, the median fold-change increase in p-tau217 between non-AD and AD participants was 6.7 (Supplementary Figure 2a). Using the ALZpath assay, with AD classification using the CSF “amyloid only” definition, the median fold-change increase in p-tau217 between non-AD and AD participants of 4.2 (Supplementary Figure 2b). Median fold changes were similar within each assay when using the CSF “amyloid and p-tau” definition (Supplementary Figure 2c,d) or the “clinical AD status” definition (Supplementary Figure 2e,f).

#### 3.2.2 Amyloid PET cohort

Plasma p-tau217 measurements correlated well between the Lumipulse and ALZpath assays (*rho* 0.84, p<0.001) (Supplementary Figure 1b).

Evaluating p-tau217 measurements according to AD status (non-AD or AD) classified by amyloid PET visual read, the median fold-change increase in Lumipulse p-tau217 between non-AD and AD participants was 3.9 (Supplementary Figure 3a). Using the ALZpath assay, the median fold-change increase in p-tau217 between non-AD and AD participants was 3.2 (Supplementary Figure 3b).

### 3.3 Comparison of AD status classification by p-tau217 assays

#### 3.3.1 CSF cohort

In the CSF cohort, ROC analysis for prediction of AD status based on the CSF “amyloid only” definition, using only age and sex as the predictors, gave an AUC of 0.673 (95% CI 0.605-0.741). Lumipulse p-tau217 on its own gave an AUC of 0.947 (95% CI 0.919-0.974, DeLong test *p* < 0.0001 compared with model including age and sex alone). A combined model including age, sex and Lumipulse p-tau217 gave an AUC of 0.950 (95% CI 0.925-0.974, not significantly different to Lumipulse p-tau217 alone). ALZpath p-tau217 on its own gave an AUC of 0.940 (95% CI 0.914-0.967, DeLong test *p* < 0.0001 compared with model including age and sex alone). A combined model including age, sex and ALZpath p-tau217 gave an AUC of 0.946 (95% CI 0.921-0.970, not significantly different to ALZpath p-tau217 alone) (Table 2). In individuals with age, sex, serum creatinine and BMI data available (n = 40), adding these covariates to the plasma p-tau217 models did not alter the result (Supplementary Table 2).

**Table 2:** ROC analyses: areas under the curve with adjustments for covariates for the CSF and amyloid PET cohorts.

|  | CSF cohort, n = 257 |  |  | Amyloid PET cohort, n = 76 |  |  |
| --- | --- | --- | --- | --- | --- | --- |
| Model | AUC | Lower<br>95% CI | Upper<br>95% CI | AUC | Lower<br>95% CI | Upper<br>95% CI |
|  | *AD status defined by CSF*<br>*$A\beta_{42}/A\beta_{40}$ only* |  |  | *AD status defined by amyloid*<br>*PET visual read* |  |  |
| Age + Sex | 0.673 | 0.605 | 0.741 | 0.547 | 0.409 | 0.686 |
| Lumipulse p-tau217 | 0.947<sup>a</sup> | 0.919 | 0.974 | 0.879 | 0.778 | 0.980 |
| ALZpath p-tau217 | 0.940<sup>a</sup> | 0.914 | 0.967 | 0.880 | 0.786 | 0.973 |
| Age + Sex +Lumipulse<br>p-tau217 | 0.950<sup>a</sup> | 0.925 | 0.974 | 0.877 | 0.775 | 0.978 |
| Age + Sex + ALZpath p-tau217 | 0.946<sup>a</sup> | 0.921 | 0.970 | 0.887 | 0.796 | 0.978 |

Note: <sup>a</sup> $p < 0.001$ (DeLong test) relative to model including age and sex alone. Abbreviations: amyloid-beta 42 to amyloid-beta 40 ratio ( $A\beta_{42}/A\beta_{40}$ ), Alzheimer's disease (AD), area under the receiver operative characteristics curve (AUC), confidence interval (CI).

Using the CSF “amyloid and p-tau” definition, the Lumipulse p-tau217 assay on its own gave an AUC of 0.938 (95% CI 0.910-0.966), and the ALZpath p-tau217 assay on its own gave an AUC of 0.914 (95% CI 0.879-0.949). Using the AD status definition based on “clinical AD status”, the Lumipulse p-tau217 assay on its own gave an AUC of 0.948 (95% CI 0.923-0.973), and the ALZpath p-tau217 assay on its own gave an AUC of 0.926 (95% CI 0.894-0.958). Combined models including age, sex, serum creatinine and BMI are detailed in Supplementary Table 3.

#### 3.3.2 Amyloid PET cohort

In the amyloid PET cohort, ROC analysis for prediction of AD status based on amyloid PET visual read, using only age and sex as the predictors, gave an AUC of 0.547 (95% CI 0.409-0.686). Lumipulse p-tau217 on its own gave an AUC of 0.879 (95% CI 0.778-0.980, DeLong test *p* < 0.0001 compared with a model including age and sex alone). A combined model including age, sex and Lumipulse p-tau217 gave an AUC of 0.877 (95% CI 0.775-0.978, not significantly different compared to AUC from model including Lumipulse p-tau217 alone); further including serum creatinine (n = 46) did not alter the result (AUC 0.837, 95% CI 0.703-0.972). ALZpath p-tau217 assay on its own gave an AUC of 0.880 (95% CI 0.786-0.973). A combined model using age, sex and ALZpath p-tau217 gave an AUC of 0.887 (95% CI 0.796-0.978, not significantly different compared to AUC from model including ALZpath p-tau217 alone) (Table 2); further including serum creatinine did not alter the result (AUC 0.864, 95% CI 0.751-0.976). No statistically significant differences were identified between AD status prediction models when adding age, sex, or serum creatinine to the plasma p-tau217 assay models (Supplementary Table 4).

### 3.4 Derivation of clinical cut-points for p-tau217

A dual cut-point approach was used and varied levels of sensitivity and specificity were applied to the data to determine p-tau217 cut-points to be used clinically at the optimal highest sensitivity and specificity percentage to achieve fewer than 20% of individuals in the intermediate zone and greater than 90% sensitivity and specificity [31].

#### 3.4.1 CSF cohort

When AD status was classified using the CSF amyloid only definition, using the Lumipulse p-tau217 assay, the cut-points at 95% sensitivity and 95% specificity, were 0.150 and 0.380 pg/mL. The percentage of individuals in the intermediate zone between these cut-points was 18.7% (Supplementary Table 5). Within this cohort the negative predictive value (NPV) of the lower cut-point was 90.2% (95% CI 81.7-95.7) and the positive predictive value (PPV) of the upper cut-point was 96.1% (95% CI 91.1-98.7), achieving an overall accuracy of the test for individuals receiving a definitive classification of low (non-AD) or high (AD) p-tau217 of 93.8% (Supplementary Table 6). To further optimize the cut-points, we investigated options that fixed the sensitivity at 95% and increased the specificity while keeping the proportion in the intermediate zone at <20%. The optimal cut-point pair was 0.153 and 0.422 pg/mL (97% specificity), which gave 19.4% of individuals in the intermediate zone, an NPV of 90.4% (95% CI 80.6-95.0) and a PPV of 97.6% (95% CI 93.2-99.5), with an overall accuracy for individuals receiving a definitive classification of low (non-AD) or high (AD) p-tau217 of 94.7% (Table 3).

**Table 3:** Performance of 95% sensitivity and maximized specificity cut-points in the CSF derivation cohort (AD status defined by CSF amyloid only definition) and amyloid PET validation cohort.

|  |  |  |  |  | CSF cohort (derivation) n = 257 |  |  |  | Amyloid PET cohort (validation) n = 76 |  |  |  |
| --- | --- | --- | --- | --- | --- | --- | --- | --- | --- | --- | --- | --- |
| Plasma p-tau217 assay | Sensitivity % | Specificity % | Lower cut-point | Upper cut-point | % in Intermediate zone | Negative predictive value of lower cut-point (95% CI) | Positive predictive value of upper cut-point (95% CI) | Accuracy of test for individuals receiving a definite classification of low or high | % in Intermediate zone | Negative predictive value of lower cut-point (95% CI) | Positive predictive value of upper cut-point (95% CI) | Accuracy of test for individuals receiving a definite classification of low or high |
| Lumipulse | 95.0 | 97.0 | 0.153 | 0.422 | 19.4 | 90.4<br>(80.6-95.0) | 97.6<br>(93.2-99.5) | 94.7 | 34.2 | 88.9<br>(65.3-98.6) | 90.6<br>(75.0-98.0) | 90.0 |
| ALZpath | 95.0 | 95.0 | 0.220 | 0.486 | 23.4 | 87.1<br>(76.2-94.3) | 96.3<br>(91.6-98.8) | 93.4 | 26.3 | 85.7<br>(42.1-99.6) | 87.8<br>(75.2-95.4) | 87.5 |

Abbreviations: Alzheimer's disease (AD), confidence interval (CI).

Using the ALZpath p-tau217 assay, the cut-points at 95% sensitivity and 95% specificity were 0.220 and 0.486 pg/mL, the percentage of individuals in the intermediate zone between these cut-points was 23.4%, NPV of the lower cut-point was 87.1% (95% CI 76.2-94.3), the PPV of the upper cut-point was 96.3% (95% CI 91.6-98.8), and overall accuracy of the test for individuals receiving a definitive classification of low (non-AD) or high (AD) p-tau217 was 93.4% (Table 3).

Cut-points at varied levels of sensitivity and specificity and related metrics were also calculated in comparison to AD status classified using the CSF “amyloid and p-tau” definition or the “clinical AD status” definition. Both these definitions gave wider intermediate zones overall than the cut-points derived using the “CSF amyloid only definition” (Supplementary Table 5) but the NPV, PPV and accuracy for both assays remained above 90% regardless of the definition used when employing the respective 95% sensitivity and 95% specificity cut-points (Supplementary Table 6).

Confusion matrices showing the numbers of participants in the categories True CSF Positive and True CSF Negative, according to plasma categorizations of Low, Indeterminate and high p-tau217 utilizing the 95% sensitivity and 95% specificity cut-points for both assays, showed that for the Lumipulse assay, those in the indeterminate plasma p-tau217 category were overall still more likely to be true CSF positive, whereas for the ALZpath assay the distribution depended on the particular CSF reference used (Supplementary Table 7a-c).

#### 3.4.2 Amyloid PET cohort

To validate the optimal p-tau217 cut-points determined from the CSF cohort (95% sensitivity and 97% specificity cut-points of 0.153 pg/mL and 0.4222 pg/mL), we next applied these cut-points to the amyloid PET cohort, the resulting percentage of individuals in the intermediate zone was 34.2%, the NPV of the lower cut-point was 88.9% (95% CI 65.3-98.6) and the PPV of the upper cut-point was 90.6% (95% CI 75.0-98.0), achieving an overall accuracy of the test for individuals receiving a definitive classification of low (non-AD) or high (AD) p-tau217 of 90.0%. The CSF cohort cut-points and related metrics for both the Lumipulse and ALZpath assays applied to the amyloid PET cohort are detailed in Table 3.

Within-cohort derivation of cut-points and related metrics for the amyloid PET cohort was also undertaken (Supplementary table 8) and this gave large proportions within the intermediate zones (of 73.7% for Lumipulse and 43.4% for ALZpath assays utilizing the within-cohort 95% sensitivity and 95% specificity cut-points for each assay respectively), with confusion matrices showing that the majority of individuals with indeterminate results were amyloid PET positive (Supplementary table 7d,e). In contrast, application to the amyloid PET cohort of cut-points derived from the CSF cohort when AD status was defined using the CSF “amyloid and p-tau” or the “clinical AD status” definitions, gave intermediate ranges and related metrics comparable to those obtained using the cut-points derived using the CSF “amyloid only” definition (Supplementary Tables 8 and 7f-k).

### 3.5 Effects of renal function impairment on plasma p-tau217

To assess the effects of impaired renal function on plasma p-tau217 measurements, we analyzed plasma from a cohort of 58 cognitively normal (CN) patients at chronic kidney disease (CKD) stages 1-4 using the Lumipulse p-tau217 assay. We applied the cut-points from the CSF cohort (0.153 and 0.422 pg/mL), and compared the CN-CKD samples to the CSF cohort samples classified as AD and non-AD. Three (i.e. 5.1%) of the CN-CKD stage 1-4 samples tested above the upper cut-point of 0.422pg/mL and 25 samples (i.e. 43.1%) tested in the intermediate zone between 0.153 and 0.422 pg/mL. The median (IQR) p-tau217 concentrations were as follows: for the AD samples 0.727 (0.437-1.068) pg/mL, non-AD samples 0.108 (0.061) pg/mL (p-value <0.001 when compared to AD), CKD-1 0.119 (0.082-0.190) pg/mL, CKD-2 0.134 (0.106-0.205) pg/mL, CKD-3a 0.164 (0.132-0.218) pg/mL, CKD-3b 0.208 (0.136-0.257) pg/mL, and CKD-4 0.213 (0.145-0.261) pg/mL (Figure 1). There was no statistically significant association between p-tau217 and CN-CKD (either as an ordinal or as a binary for CKD 3a and above) after adjustment for age, sex, or BMI where data were available (Supplementary Table 9).

**Figure 1:**
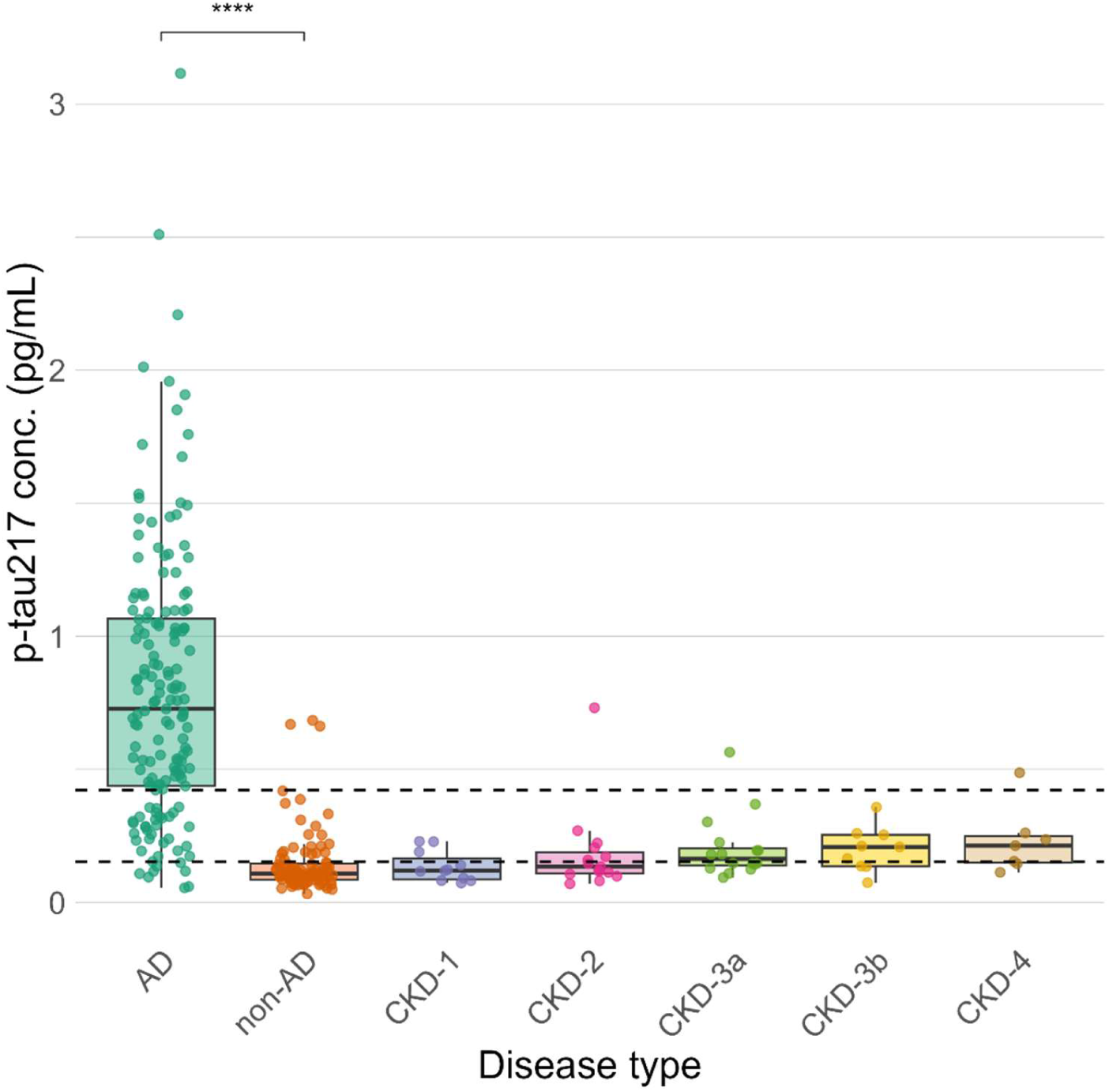
Lumipulse plasma p-tau217 measurements in renal function impairment samples (from the cognitively normal-chronic kidney disease (CN-CKD) cohort including samples at CKD stage 1 (n=11), stage 2 (n=15), stage 3a (n=16), stage 3b (n=9), stage 4 (n=7)) compared to Alzheimer’s disease (AD) and non-AD samples (from the CSF cohort with AD (n=159), and non-AD (n=98), box plots show median ± IQR; dotted lines represent the 0.153 and 0.422pg/mL p-tau217 cut-points derived from the CSF cohort, p<0.001 **** using a Wilcoxon signed-rank test).

### 3.6 Pre-analytical variation

Four experiments assessing the effects of varied pre-analytical sampling conditions on relative plasma p-tau217 concentrations were undertaken using both the Lumipulse and ALZpath assays. For the Lumipulse assay, Experiments 1 (pre-centrifugation delay at room temperature, Figure 2a), 2 (pre-centrifugation delay at 2-8°C, Figure 2b) and 4 (number of freeze-thaw cycles, Figure 2d) did not yield statistically significant differences (p>0.05, Supplementary Table 10) between groups. In Experiment 3 (post-centrifugation delay, Figure 2c), a significant difference was observed between post-centrifugation delay groups (p = 0.0047, Supplementary Table 10), which post-hoc testing revealed to be between the post-centrifuged samples stored at 4 and 24 hours in 2-8°C compared to the samples stored for 2 weeks at −20°C (median relative change at 4hr, 2-8°C = 0.96; at 24hr, 2-8°C = 0.94; at 2 weeks, −20°C = 1.04, Supplementary Table 10). This finding indicates that, using the Lumipulse assay, a transient subtle reduction of ∼5% in p-tau217 level was seen when post-centrifugation samples were kept in the fridge between 4 to 24 hours, which could be mitigated by storing at −20°C instead, for up to 2 weeks.

**Figure 2:**
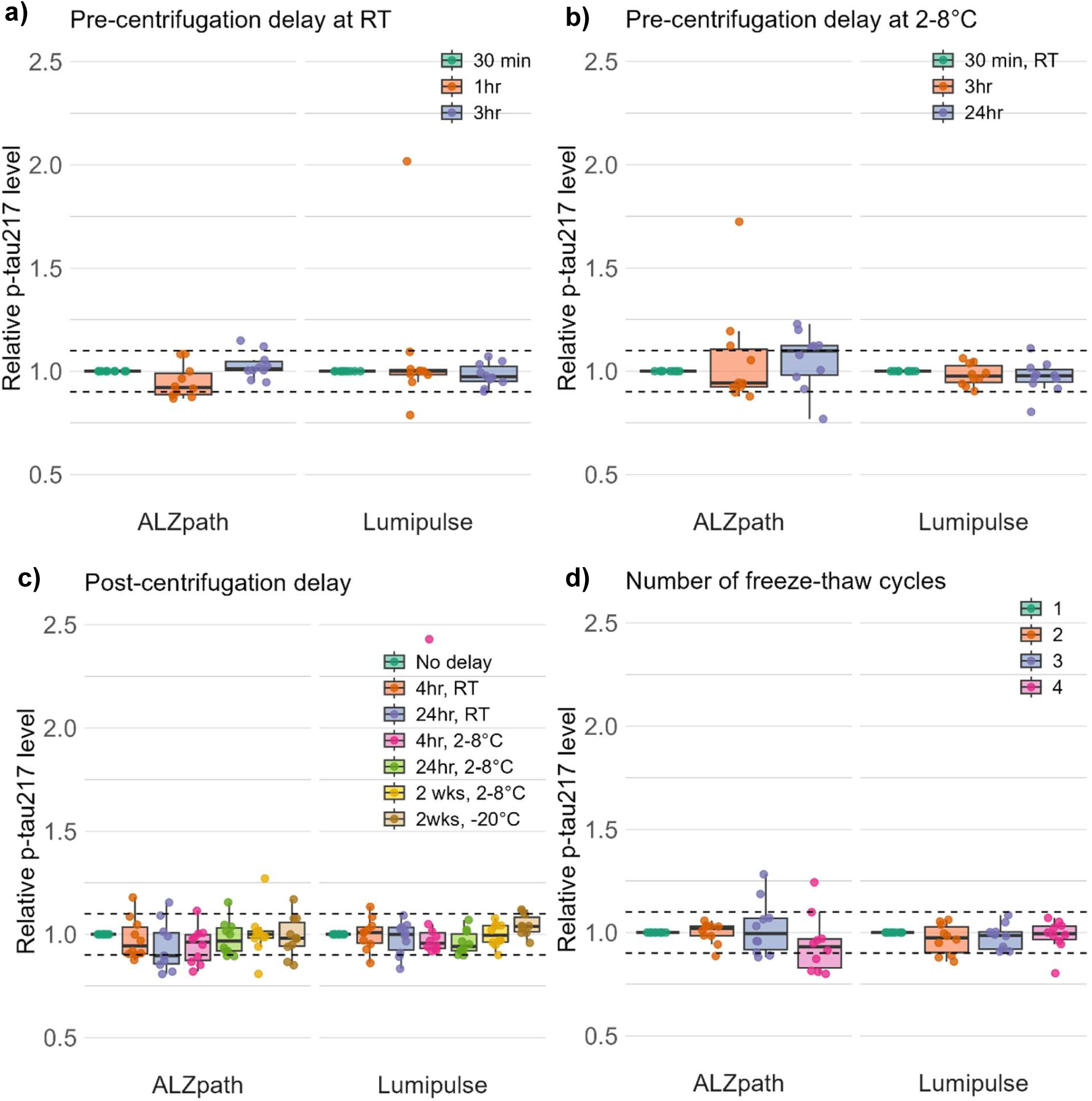
Relative change in concentration of plasma p-tau217 across pre-analytical sample handling conditions: **a)** pre-centrifugation delay at RT, **b)** pre-centrifugation delay at 2-8°C, **c)** post-centrifugation delay, **d)** number of freeze-thaw cycles, compared to the baseline condition using the ALZpath and Lumipulse assays (n = 10 per experiment, data presented as median ± IQR, dotted lines represent the lower and upper bounds of a 10% difference threshold from the baseline condition).

Using the ALZpath assay, Experiment 1 (pre-centrifugation delays of samples in room temperature, Figure 2a) led to subtle but statistically significant reductions in relative p-tau217 levels (median relative level at 1 hour = 0.92, Friedman test p = 0.007). Post-hoc testing revealed that this difference occurred between 1 and 3 hours of pre-centrifugation delay (Supplementary Table 10). No difference existed between the base condition and 3-hour time point, indicating normalization of plasma p-tau217 levels with longer pre-centrifugation delays at room temperature. Plasma p-tau217 using the ALZpath assay was also robust to pre-analytical handling in Experiments 2, 3, and 4 (Supplementary Table 10).

Across all four experiments, the median relative change across conditions did not exceed the ±10% threshold considered likely to be clinically significant (Figure 2). We noted individual-level variability in relative p-tau217 changes (for example, one outlier in the Lumipulse assay at the 1h time point in Experiment 1) which could not be explained by a low absolute p-tau217 level (Supplementary Table 10).

### 3.7 Lumipulse plasma p-tau217 assay lot-to-lot variability

A comparison of plasma p-tau217 measurements from 4 kit lots of the Lumipulse plasma p-tau217 assay kits (Fujirebio) (two version 1 lots: 4128 and 5066; two version 2 lots 5084 and 5086) was carried out to assess lot-to-lot variability in measurements in a subset (n = 30) of samples from the CSF cohort. We reported strong correlations (*R*>0.95) in plasma p-tau217 measurements between all 4 kit lots compared to each other (p<0.001) indicating high concordance of plasma p-tau217 results across assay kit lots (Supplementary Figure 4). Three out of 30 individuals had discordant classifications across lots relative to the established dual cut-points; discordances did not occur due to the same lot. Two of these were re-classifications across the lower cut-point and one was across the upper cut-point.

### 3.8 Plasma sample transport temperature comparison

In a small set of 10 samples collected from the ongoing amyloid PET cohort, an excellent correlation was observed between Lumipulse p-tau217 measurements from samples stored and transported at −20°C and −80°C (R = 0.998, Supplementary Figure 5) indicating that intermediate storage at −20°C and transport in a Bio-freeze device at −20 to 0°C would allow for robust measurement of plasma p-tau217 in the ADAPT stage 3 trial.

## 2. Discussion

For stage 1 of the ADAPT study, we aimed to derive and validate plasma p-tau217 cut-points for clinical use. We evaluated plasma p-tau217 measurements by two commercially available assays against two independent cohorts with different gold-standard biomarkers used clinically for AD diagnosis (CSF AD biomarkers and amyloid PET).

Though measurements by the two assays correlated well within each cohort, the Lumipulse assay performed better in terms of median fold change compared with the ALZpath assay in both cohorts, and this is consistent with the reports of a recent plasma p-tau round-robin study [6].

Whilst ROC analyses showed overall similar AUC for both assays within each cohort, the Lumipulse assay was superior in terms of giving a lower percentage of individuals with indeterminate results when applying 95% sensitivity and maximized specificity cut-points (0.153/0.422pg/mL). Of note, AUC were lower within the amyloid PET cohort, and applying the CSF-cohort derived cut-points for either assay to the amyloid PET cohort resulted in the proportion of individuals with indeterminate results exceeding 20%. Previous studies have suggested that there can be discordance between amyloid PET visual reads and quantitative measures in 5-14% of cases, particularly in cases with borderline values [32, 33], which could explain the lower concordance identified here. It is also possible that despite having similar prevalence of AD pathology overall, the two cohorts had differing degrees of amyloid and/or tau burden; in the amyloid PET cohort 51.3% had significant amyloid burden and 11.8% had mild amyloid burden, but in the CSF cohort the degree of amyloid burden was not known.

Our investigation of pre-analytical sample handling factors showed that plasma p-tau217 measured by either the Lumipulse or ALZpath assays is stable across a range of pre-analytical handling variations, with any small statistically significant differences in individual conditions not exceeding 10% relative change. These findings align with and extend prior studies [28, 34, 35], showing p-tau217 stability under conditions such as delayed centrifugation, refrigeration, and freeze-thaw cycles. Notably, p-tau217 remained stable when stored post-centrifugation for up to two weeks at 2-8°C or −20°C (−20°C showed preferable higher stability), supporting its feasibility for use in remote settings. Further to this, we showed that post-centrifugation storage of plasma samples at −20°C for up to two weeks followed by transport at −20-0°C maintained this stability of p-tau217 compared to −80°C post-centrifugation storage and transport on dry ice. Based on these findings, a recommended sample handling protocol would include:

- Blood can be stored for up to 3 hours at room temperature or 24 hours refrigerated before centrifugation.
- Plasma can be stored for up to 2 weeks frozen at −20°C (or refrigerated at 2-8°C if necessary).
- Up to four freeze-thaw cycles prior to p-tau217 measurements are acceptable.
- Plasma can be transported at sub-zero temperature using a temperature-controlled transport system (e.g. Bio-Freeze) without the need for dry ice.
- The robustness of plasma p-tau217 measurement to these variations indicates that it is suitable to apply in settings where immediate long term storage at −80°C is not available, and therefore may allow for its use to be extended to settings in which currently CSF testing (which requires more stringent sample handling due to susceptibility of amyloid peptides to pre-analytical variation) is not available.

This study has some limitations. Ethnicity was not systematically recorded and patients from Black, Asian and other ethnicities comprised 11% of the CSF cohort and 25% of the amyloid PET cohort. While inter-ethnic differences in plasma p-tau17 levels have been noted in other studies [36], the results are mixed, and there is no clear indication that differing cut-points would be appropriate to apply based on ethnicity. Further research should be carried out with the inclusion of larger numbers of patients from ethnically diverse backgrounds, to determine any differences in the accuracy of using plasma p-tau217 to diagnose AD. Another limitation of is the partial availability of data on participant comorbidities in the original derivation and validation cohorts [21]. We addressed this limitation by analyzing plasma p-tau217 in a separate cohort of patients with ADPKD and CKD stages 1-4 aged under 60 years. This cohort did not have a reference standard amyloid biomarker (CSF or PET) but the background prevalence of amyloid pathology was likely to be <20% according to age [27], and although we found that plasma p-tau217 was not frequently elevated in CKD to levels that could be mistaken with an AD diagnosis, CKD was associated with elevation of plasma p-tau217 predominantly into the intermediate zone. We therefore advocate that serum creatinine levels should be assessed for patients in whom plasma p-tau217 is being used as an adjunct to diagnosis, and extra caution should be taken when interpreting plasma p-tau217 results in patients with CKD [37, 38].

Recently the US Federal Drug Administration approved the Lumipulse G p-tau217/Aβ_42_ ratio as a plasma test to aid in Alzheimer’s disease diagnosis in symptomatic patients aged 55 years and older. We did not test plasma Aβ_42_ in the ADAPT stage 1 study as the literature at the time of study design was in support of utilizing p-tau217 as a single biomarker in this context. Subsequent new data indicate that incorporating plasma Aβ_42_ in a ratio with p-tau217 does not substantially change the percentage of individuals with indeterminate results relative to using plasma p-tau217 as a single biomarker [39]. The susceptibility of plasma Aβ_42_ to pre-analytical handling variation [22, 40] may also make the p-tau217/Aβ_42_ ratio more difficult to apply in clinical routine, hence we have implemented the Lumipulse p-tau217 assay in our clinical laboratory on its own rather than using this ratio.

In conclusion, here we present cut-points (0.153/0.422pg/mL), derived against CSF amyloid status classification and validated against amyloid PET, to be used clinically for the interpretation of Lumipulse (Fujirebio) plasma p-tau217 results. The Lumipulse (Fujirebio) plasma p-tau217 assay consistently showed higher performance than the ALZpath (Quanterix) assay when employing a dual cut-point interpretation, in terms of having both higher median fold change between AD and non-AD groups, and lower proportion of cases with indeterminate results. The lower and upper cut-points derived at 95% sensitivity and 97% specificity using the Lumipulse assay will be used as the clinical cut-points for interpretation of plasma p-tau217 measured from samples sent from across the UK at the UKAS-accredited Neuroimmunology and CSF Laboratory (NHS) which is the reference laboratory for dementia biomarkers in the UK. The Lumipulse assay will be implemented in stage 3 of the ADAPT study: a randomized controlled clinical trial of result disclosure in community memory clinics. The observations on stability of plasma p-tau17 to variations in the pre-analytical handling factors assessed in our study have been used to inform the sample handling protocol that will be sent to trial sites. Robustness of sample measurements across different kit lots and assay versions indicates that the derived cut-points are likely to be stable in clinical routine testing and across the trial’s recruitment period, but we also plan to confirm this by assessing the performance of the cut-points over a three-year period in relation to CSF and amyloid PET.

## Acknowledgements

AK and JMS conceived the study. KW, RW, IG-B and KT performed the experiments. MH and AJH supervised the experiments. KW, RW, MCBD and OS-A curated the data. AK, KW and RW analyzed the data. AK, KW, RW and JMS drafted the initial manuscript. KW and RW drafted the figures. All authors reviewed and edited the manuscript.

We are very grateful to participants in the UCL DRC Wolfson CSF study and Biomarkers and Genetics in Cognitive disorders cohorts, the Imperial College London pAPC study cohort and the PKD clinical genetics clinic cohort sponsored by the PKD Charity at the Royal Free Hospital.

We thank Floey Urban, Boglarka Zilahi and Millie Beament for their efforts in the collection of samples for the Biomarkers and Genetics in Cognitive Disorders study, and Frankie O’Shea and fellows at the UCL DRC for their efforts in collection of samples for the Wolfson CSF study. We are grateful to Fotini Mastorakou, Liam O’Donohue and Miles Chapman at the Neuroimmunology and CSF laboratory.

## Sources of Funding

The ADAPT study is funded by the Blood Biomarker Challenge grant ARUK-BBC2023-002 (funding partners Alzheimer’s Society, Alzheimer’s Research UK, Postcode Innovation Trust, People’s Postcode Lottery, the National Institute for Health and Social Care Research),

AK, MH, MPL, NCF, HZ and JMS acknowledge the support of the UCLH NIHR Biomedical Research Centre.

The Fluid Biomarker Laboratory at UCL is supported by the UK Dementia Research Institute (UKDRI-1003) and the National Institute for Health and Care Research University College London Hospitals Biomedical Research Centre.

## Disclosures

### Consent statement

All human subjects provided written informed consent.

### Data availability statement

The anonymized data that support the findings of this study are available on request from qualified academic investigators, after approval of a proposal and with a signed data access agreement. Data will be shared for the sole purpose of replicating procedures and results. Requests should be directed to the corresponding author

