## Supplementary materials for "The Alzheimer’s Disease Diagnosis and Plasma Phospho-Tau217 (ADAPT) study stage 1: validating clinical cut-points against CSF and amyloid PET"

### Supplementary Figures

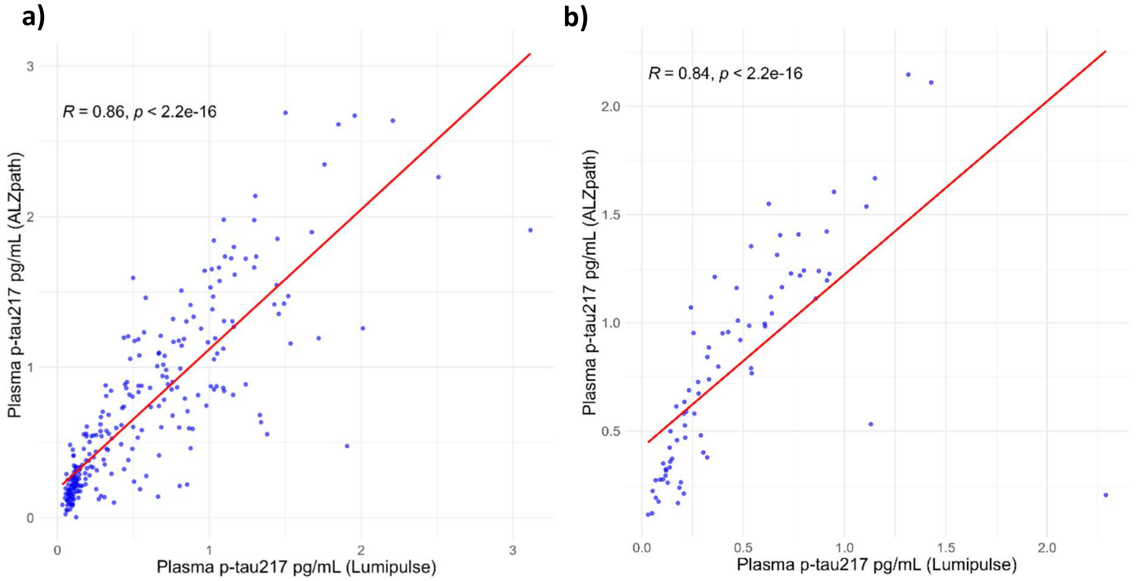


**Supplementary Figure 1:** Correlation between Lumipulse and ALZpath assay p-tau217 measurements with Spearman's rho (R) correlation coefficient and p-value for the **a)** CSF cohort (n=257) and **b)** amyloid PET cohort (n=76).

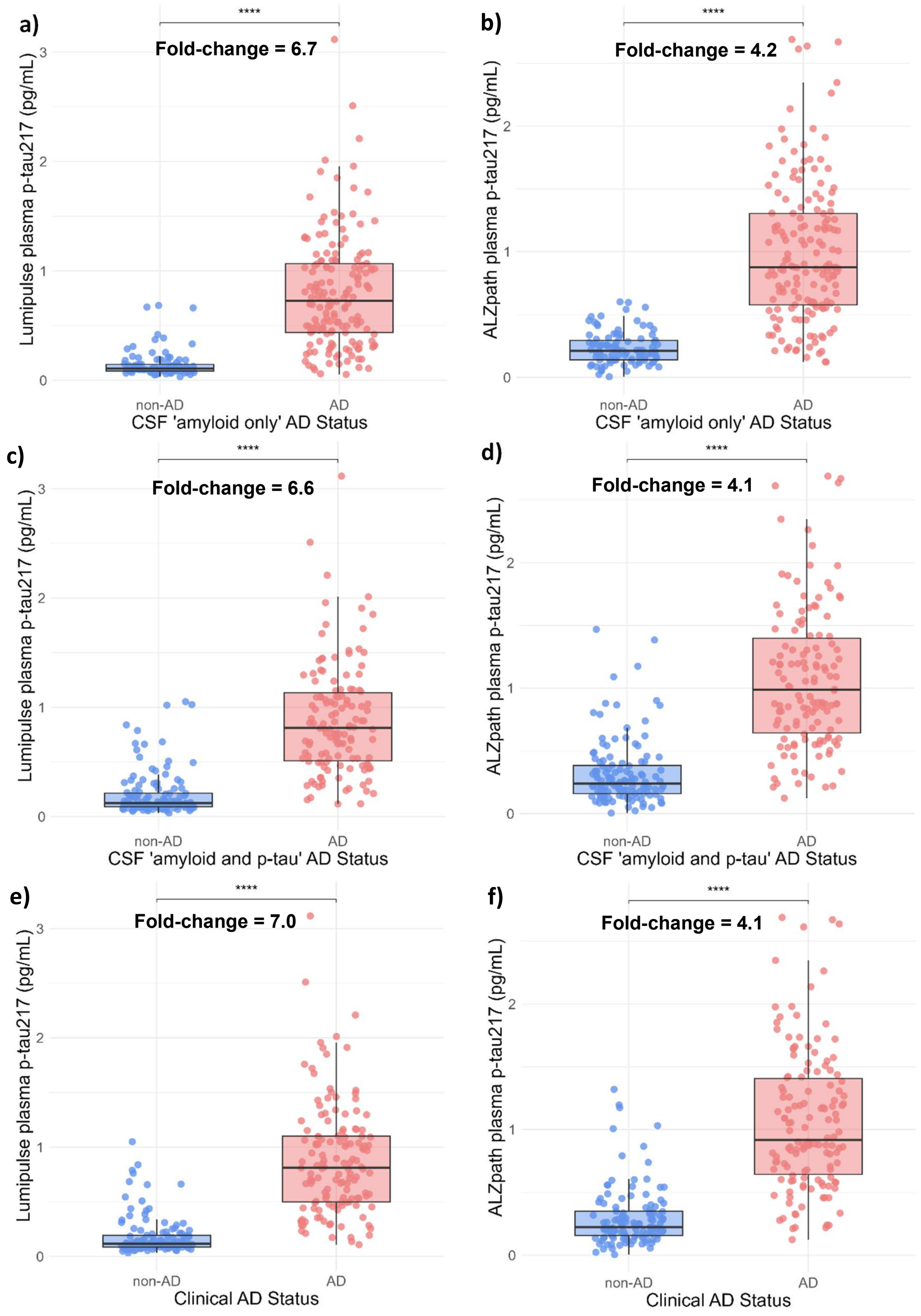


**Supplementary Figure 2:** Plasma p-tau217 measurements in patients classified using gold-standard biomarkers as non-Alzheimer's disease (non-AD) and AD in the CSF cohort via varied classification methods of: CSF $A\beta_{42}/A\beta_{40}$ ratio only using the **a)** Lumipulse and **b)** ALZpath assays; CSF $A\beta_{42}/A\beta_{40}$ ratio and p-tau181 using the **c)** Lumipulse and **d)** ALZpath assays**;** and most recent clinical diagnosis (informed by CSF) using the **e)** Lumipulse and **f)** ALZpath assays (n = 257, box plot shows median ± IQR, median fold-change in plasma p-tau217 between non-AD and AD patients indicated on plots, $p<0.001$ **** using a Wilcoxon signed-rank test).

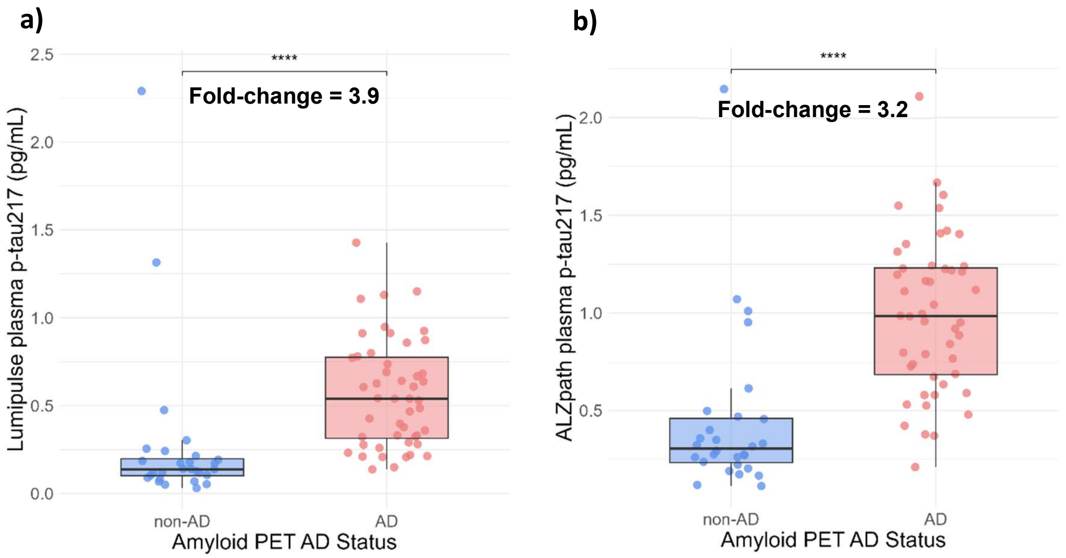


**Supplementary Figure 3:** Plasma p-tau217 measurements in patients classified using an amyloid PET visual read as an independent gold-standard biomarker as non-Alzheimer's disease (non-AD) and AD in the amyloid PET cohort using the **a)** Lumipulse and **b)** ALZpath assays (n = 76, box plot shows median ± IQR, median fold-change in plasma p-tau217 between non-AD and AD patients indicated on plots, p<0.001 **** using a Wilcoxon signed-rank test).

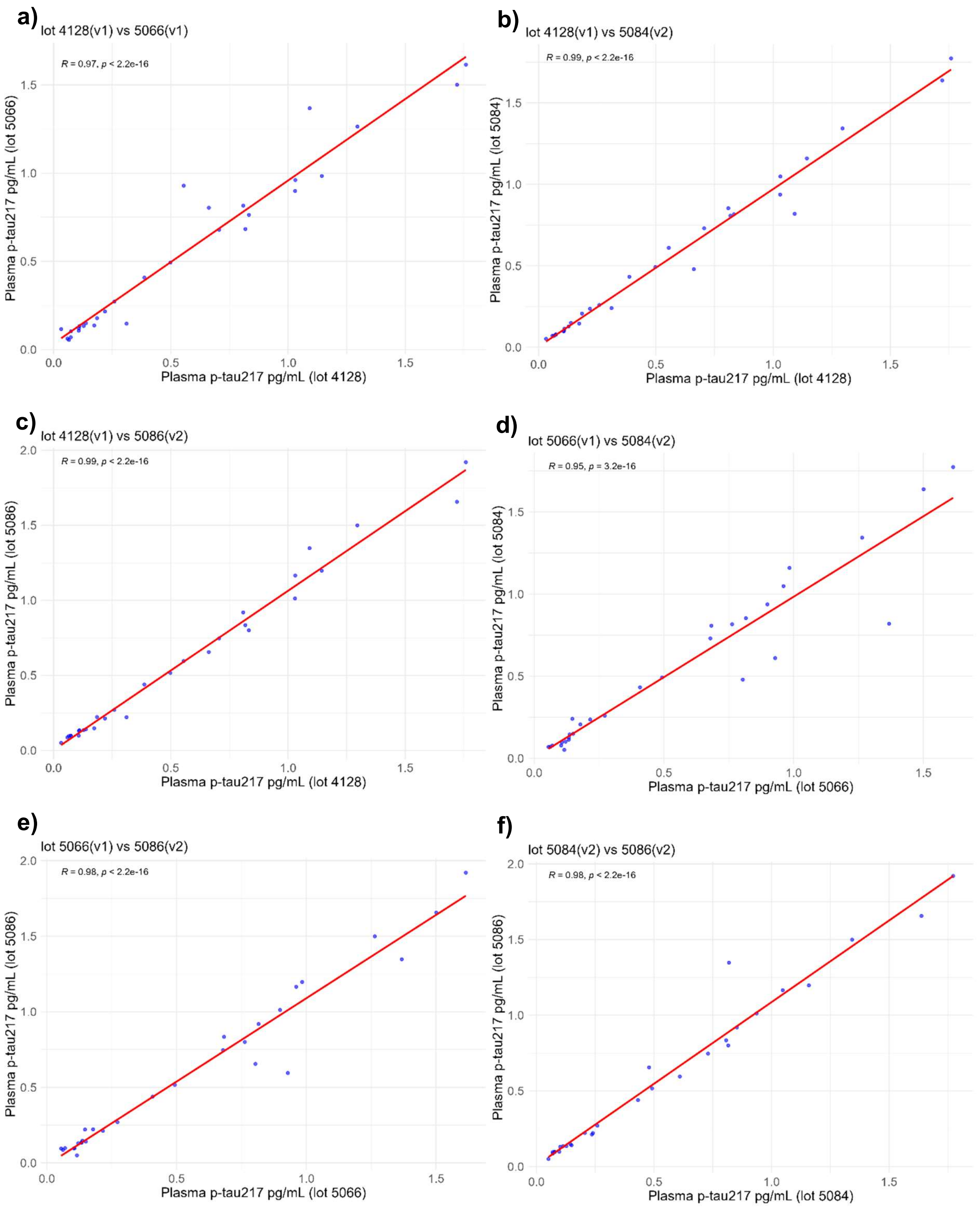


**Supplementary Figure 4:** Plasma p-tau217 measurement correlations using 4 different Lumipulse G plasma p-tau217 (Fujirebio) assay kit lots (from version 1 (v1) and version 2 (v2)): **a)** lot 4128 (v1) vs lot 5066 (v1), **b)** lot 4128 (v1) vs lot 5084 (v2), **c)** lot 4128 (v1) vs lot 5086 (v2), **d)** lot 5066 (v1) vs lot 5084 (v2), **e)** lot 5066 (v1) vs lot 5086 (v2), and **f)** lot 5084 (v2) vs lot 5086 (v2), in a subset of plasma samples selected from the CSF cohort (n = 30). Correlation between lots reported using Spearman's *rho* (R) ($p<0.001$ for all lot comparisons).

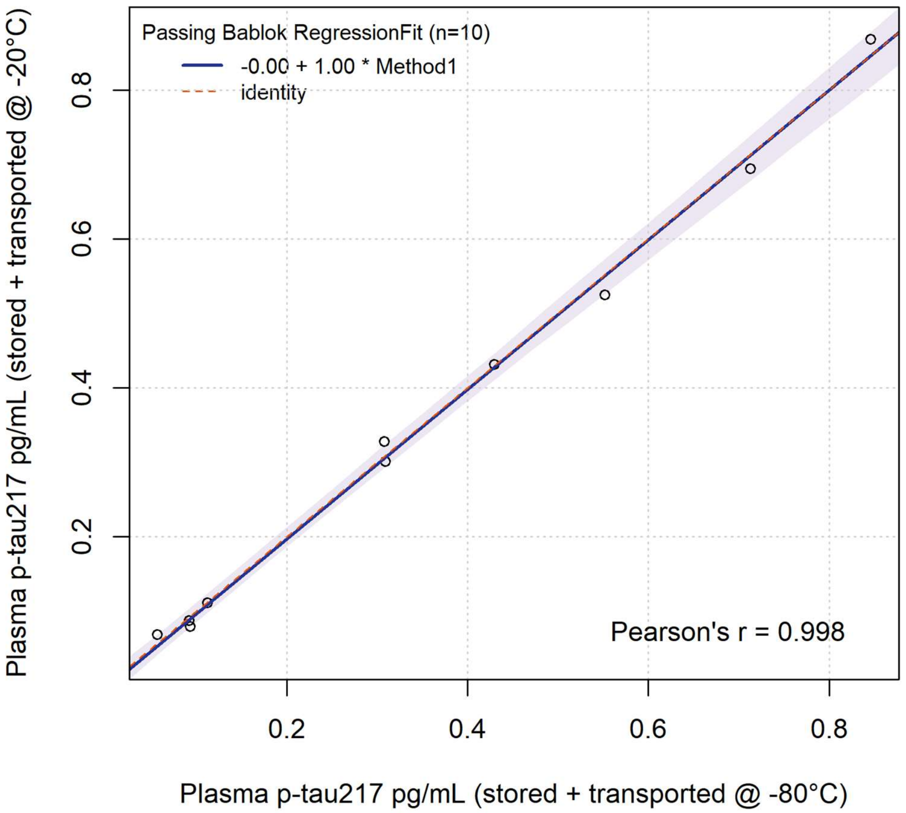


**Supplementary Figure 5:** Passing Bablok regression fit for plasma p-tau217 measurements (pg/mL) in samples from the ongoing amyloid PET cohort (n = 10) that either had: intermediate storage of centrifuged plasma aliquots at -80°C followed by transport on dry ice for <24 hrs and storage at -80°C until analysis (Method 1), or intermediate storage of centrifuged plasma aliquots at -20°C followed by Bio-freeze transport at -20-0°C for <24 hrs and storage at -80°C until analysis (Method 2). Shading indicates the 95% confidence bounds (calculated using the bootstrap(quantile) method).

### Supplementary Tables

**Supplementary Table 1:** CKD cohort participant demographic characteristics.

| **Variable** | **CKD cohort, n = 58** |
|---|---|
| Age at sampling (years): mean (SD) | 44.4 (8.5) |
| Sex (male): n (%) | 32 (55.2) |
| Ethnicity: n (%) | |
| White | 27 (46.5) |
| Black | 3 (5.2) |
| Asian | 3 (5.2) |
| Other | 6 (10.4) |
| Not stated/known | 19 (32.8) |
| CKD stage: n (%) | |
| G1 | 11 (19.0) |
| G2 | 15 (25.9) |
| G3a | 16 (27.6) |
| G3b | 9 (15.5) |
| G4 | 7 (12.1) |
| BMI: median (IQR) | 25.0 (24.0-28.8), n = 40 (69.0%) |

*Abbreviations: chronic kidney disease (CKD), body mass index (BMI), interquartile range (IQR), standard deviation (SD).*

**Supplementary Table 2:** Multiple logistic regression coefficients (with 95% CI and p-value significance levels) for varied models of AD status prediction (using the CSF “amyloid only” definition) of age, sex, serum creatinine, and BMI on p-tau217 measurements in the CSF cohort, using the Lumipulse and ALZpath assays, used to predict ROC AUCs. The subset refers to the group in which all variables were available.

| Model | Coefficient (95% CI; p-value) | | | | | AIC |
|---|---|---|---|---|---|---|
| | **Age** | **Sex** | **SCr** | **BMI** | **p-tau217** | |
| **Model 1** (no plasma p-tau217, n=257) | 0.047 (0.011, 0.084; 0.012) * | -1.126 (-1.701, -0.576; <0.001) **** | - | - | - | - |
| **Model 2** Subset (no plasma p-tau217, n=40) | 0.038 (-0.036, 0.120; 0.322) | -0.535 (-1.874, 0.748; 0.419) | - | - | | 59.1 |
| **Lumipulse** | | | | | | |
| **Model 3** (n=257) | 0.091 (0.033, 0.154; 0.003) *** | -0.268 (-1.202 0.683; 0.574) | - | - | 10.149 (7.564, 13.350; <0.001) **** | - |
| **Model 4** (n=62) | 0.027 (-0.108, 0.175; 0.701) | -0.152 ( -2.596, 2.309; 0.899) | -0.026 ( -0.095, 0.015; 0.347) | - | 16.452 (7.961, 9.779; 0.003) *** | - |
| **Model 5** (Subset, n=40) | -0.074 (-0.250, 0.076; 0.338) | 1.872 ( -1.341, 6.824; 0.322) | - | - | 27.019 (11.583, 58.428; 0.015) * | 21.2 |
| **Model 6** (Subset, n=40) | -0.054 ( -0.237, 0.113; 0.511) | 2.852 -0.851, 9.281; 0.210) | -0.024 ( -0.103, 0.017; 0.358) | - | 29.787 (11.623, 68.661; 0.029) * | 22.0 |
| **Model 7** (Subset, n=40) | -0.049 (-0.276, 0.143; 0.601) | 2.974 (-0.987 10.258; 0.234) | -0.020 (-0.093, 0.019; 0.379) | -0.163 (-0.600, 0.160; 0.353) | 28.094 (11.545, 66.995; 0.023) * | 23.1 |
| **ALZpath** | | | | | | |
| **Model 8** (n=257) | 0.062 (0.005, 0.122; 0.036) * | -0.843 (-1.763, 0.055; 0.067) | - | - | 8.838 (6.612, 11.580; <0.001) **** | - |
| **Model 9** (n=62) | 0.067 (-0.074, 0.231; 0.357) | -1.595 (-5.248, 1.248; 0.303) | -0.034 (-0.114, 0.007; 0.193) | - | 16.107 (8.236, 32.020; 0.004) *** | - |
| **Model 10** (Subset, n=40) | 0.002 (-0.153, 0.159; 0.975) | -0.278 (-4.842, 3.640; 0.887) | - | - | 13.704 (6.728, 27.684; 0.006) ** | 20.1 |

| | | | | | | |
|---|---|---|---|---|---|---|
| **Model 11** (Subset, n=40) | 0.046 (-0.125, 0.223; 0.564) | 0.940 (-3.893, 6.312; 0.674) | -0.037 (-0.140, 0.007; 0.200) | - | 16.365 (7.364, 37.997; 0.016) * | 19.5 |
| **Model 12** (Subset, n=40) | 0.019 (-0.184, 0.204; 0.828) | 2.187 (-3.037, 12.338; 0.453) | -0.040 (-0.136, 0.006; 0.167) | -0.262 (-1.143, 0.160; 0.333) | 17.057 (7.385, 46.819; 0.031) * | 20.2 |

*Note: significance levels * p < 0.05, ** p < 0.01, *** p < 0.005, **** p < 0.001. Abbreviations: Akaike information criterion (AIC), Alzheimer's disease (AD), body mass index (BMI), confidence interval (CI), receiver operating characteristic area under the curve (ROC AUC), serum creatinine (SCr).*

**Supplementary Table 3:** ROC analyses: areas under the curve (AUC) with adjustments for covariates for the CSF cohort alternate AD status definitions.

| | **CSF cohort, n = 257** | | |
|---|---|---|---|
| **Model** | **AUC** | **Lower 95% CI** | **Upper 95% CI** |
| | *AD status defined by CSF $A\beta_{42}/A\beta_{40}$ and p-tau181* | | |
| Age + Sex | 0.628 | 0.560 | 0.696 |
| Lumipulse p-tau217 | 0.938 | 0.910 | 0.966 |
| ALZpath p-tau217 | 0.914 | 0.879 | 0.949 |
| Age + Sex + Lumipulse p-tau217 | 0.938 | 0.910 | 0.966 |
| Age + Sex + ALZpath p-tau217 | 0.916 | 0.881 | 0.950 |
| Age + Sex + Serum Creatinine + BMI + Lumipulse p-tau217 | 0.997 | 0.994 | 1.000 |
| Age + Sex + Serum Creatinine +BMI + ALZpath p-tau217 | 0.989 | 0.980 | 0.997 |
| | *AD status defined by most recent clinical diagnosis (informed by CSF)* | | |
| Age + Sex | 0.640 | 0.573 | 0.707 |
| Lumipulse p-tau217 | 0.948 | 0.923 | 0.973 |
| ALZpath p-tau217 | 0.926 | 0.894 | 0.958 |
| Age + Sex + Lumipulse p-tau217 | 0.951 | 0.927 | 0.974 |
| Age + Sex + ALZpath p-tau217 | 0.930 | 0.898 | 0.961 |
| Age + Sex + Serum Creatinine + BMI + Lumipulse p-tau217 | 0.972 | 0.929 | 1.000 |
| Age + Sex + Serum Creatinine + BMI + ALZpath p-tau217 | 0.972 | 0.929 | 1.000 |

*Abbreviations: amyloid-beta 42 to amyloid-beta 40 ratio ($A\beta_{42}/A\beta_{40}$), Alzheimer's disease (AD), body mass index (BMI), confidence interval (CI), receiver operating characteristic (ROC).*

**Supplementary Table 4:** Multiple logistic regression coefficients (with 95% CI and p-value significance levels) for models of AD status prediction of age, sex, serum creatinine on p-tau217 measurements in the amyloid PET cohort, using the Lumipulse and ALZpath assays, used to predict ROC AUCs. The subset refers to the group in which all variables were available.

| **Model** | **Coefficient (95% CI; p-value)** | | | | **AIC** |
|---|---|---|---|---|---|
| | **Age** | **Sex** | **SCr** | **p-tau217** | |
| **Model 1** (no plasma p-tau217, n=76) | 0.027 (-0.028, 0.083) p=0.333 | -0.139 (-1.089, 0.811) p=0.773 | - | | - |
| **Model 2** Subset (no plasma p-tau217, n=46) | 0.015 (-0.049, 0.081) p=0.633 | -0.546 (-1.749, 0.622) p=0.363 | - | | 68.7 |
| | **Lumipulse** | | | | |
| **Model 3** (n=76) | 0.040 (-0.021, 0.106) p=0.217 | -0.103 (-1.147, 0.933) p=0.844 | - | 3.312 (1.415, 5.687) p=0.002 *** | - |
| **Model 4** (Subset, n=46) | 0.020 (-0.047, 0.092) p=0.568 | -0.240 (-1.520, 1.050) p=0.712 | - | 1.912 (0.274, 4.044) p=0.046 * | 65.2 |
| **Model 5** (Subset, n=46) | 0.021 (-0.047, 0.094) p=0.555 | -0.178 (-1.651, 1.297) p=0.810 | -0.005 (-0.063, 0.053; 0.861) | 1.909 (0.273, 4.044) p=0.046 * | 67.2 |
| | **ALZpath** | | | | |
| **Model 6** (n=76) | 0.021 (-0.046, 0.095) p=0.549 | -0.517 (-1.766, 0.659) p=0.398 | - | 3.873 (2.231, 5.924) p<0.001) **** | - |
| **Model 7** (Subset, n=46) | 0.003 (-0.069, 0.078) p=0.945 | -0.358 (-1.761, 1.010) p=0.607 | - | 2.719 (1.113, 4.746) p=0.003 *** | 57.8 |
| **Model 8** (Subset, n=46) | 0.005 (-0.067, 0.082) p=0.886 | -0.158 (-1.731, 1.387) p=0.840 | -0.018 (-0.081, 0.046) p=0.579 | 2.807 (1.153, 4.963) p=0.003 *** | 59.5 |

*Note: significance levels * p < 0.05, *** p < 0.005, **** p < 0.001. Abbreviations: Akaike information criterion (AIC), Alzheimer's disease (AD), confidence interval (CI), receiver operating characteristic area under the curve (ROC AUC), serum creatinine (SCr).*

**Supplementary Table 5:** Lumipulse and ALZpath p-tau217 cut-points and intermediate zone percentages determined by varied methods of AD status classification using varied sensitivity and specificity in the CSF cohort

| Assay | 90% Sensitivity cut-point | 90% Specificity cut-point | % in Intermediate zone | 95% Sensitivity cut-point | 95% Specificity cut-point | % Intermediate zone | 97.5% sensitivity cut-point | 97.5% specificity cut-point | % Intermediate zone |
|---|---|---|---|---|---|---|---|---|---|
| *AD status defined by CSF $A\beta_{42}/A\beta_{40}$ ratio only* | | | | | | | | | |
| **Lumipulse** | 0.236 | 0.254 | 1.2 | 0.150 | 0.380 | 18.7 | 0.109 | 0.664 | 43.2 |
| **ALZpath** | 0.325 | 0.419 | 7.0 | 0.220 | 0.486 | 23.3 | 0.190 | 0.558 | 34.6 |
| *AD status defined by CSF $A\beta_{42}/A\beta_{40}$ ratio and p-tau181* | | | | | | | | | |
| **Lumipulse** | 0.301 | 0.438 | 7.4 | 0.239 | 0.670 | 24.9 | 0.157 | 0.844 | 42.0 |
| **ALZpath** | 0.458 | 0.610 | 10.9 | 0.278 | 0.870 | 36.6 | 0.212 | 1.093 | 54.9 |
| *AD status defined by most recent clinical diagnosis (informed by CSF)* | | | | | | | | | |
| **Lumipulse** | 0.301 | 0.403 | 5.8 | 0.236 | 0.660 | 23.5 | 0.178 | 0.763 | 36.2 |
| **ALZpath** | 0.458 | 0.558 | 6.9 | 0.278 | 0.798 | 31.2 | 0.220 | 1.042 | 50.4 |

*Abbreviations: amyloid-beta 42 to amyloid-beta 40 ratio ($A\beta_{42}/A\beta_{40}$), Alzheimer's disease (AD).*

**Supplementary Table 6:** Positive and negative predictive values using 95% sensitivity and 95% specificity cut-points for the CSF cohort (AD status defined by CSF Aβ42/40 “amyloid only”, CSF Aβ42/40 + p-tau181 “amyloid and p-tau” or by most recent clinical diagnosis “clinical AD status” (informed by CSF)) and for the amyloid PET cohort.

| | 95% Sensitivity cut-point | 95% Specificity cut-point | % in Intermediate zone | Negative predictive value of lower cut-point | Positive predictive value of upper cut-point | Accuracy of test for individuals receiving a definite classification of low or high |
|---|---|---|---|---|---|---|
| **CSF cohort (CSF $A\beta_{42}/A\beta_{40}$ only) Lumipulse applied to CSF cohort** | 0.150 | 0.380 | 18.7 | 90.2 | 96.1 | 93.8 |
| **CSF cohort (CSF $A\beta_{42}/A\beta_{40}$+ p-tau181) Lumipulse applied to CSF cohort** | 0.239 | 0.670 | 24.9 | 93.3 | 93.3 | 93.3 |
| **CSF cohort (CSF $A\beta_{42}/A\beta_{40}$+ p-tau181) ALZpath applied to CSF cohort** | 0.278 | 0.870 | 36.6 | 91.4 | 92.7 | 92.0 |
| **CSF cohort (Clinical diagnosis) Lumipulse applied to CSF cohort** | 0.236 | 0.660 | 23.5 | 93.3 | 93.7 | 93.5 |
| **CSF cohort (Clinical diagnosis) ALZpath applied to CSF cohort** | 0.278 | 0.798 | 31.2 | 91.4 | 93.9 | 92.7 |
| **Amyloid PET cohort Lumipulse applied to Amyloid PET cohort** | 0.160 | 1.372 | 73.7 | 88.9 | 50.0 | 85.0 |
| **Amyloid PET cohort ALZpath applied to Amyloid PET cohort** | 0.375 | 1.091 | 43.4 | 90.5 | 95.5 | 93.0 |

*Abbreviations: amyloid-beta 42 to amyloid-beta 40 ratio ($A\beta_{42}/A\beta_{40}$), Alzheimer’s disease (AD).*

**Supplementary Table 7:** Confusion matrices of True positive, True negative, False positive and False negative rates, with numbers in intermediate zones, using the 95% sensitivity and 95% specificity cut-points by varied methods of AD status definition within the CSF and amyloid PET cohorts.

| ***a)*** *CSF cohort: AD status determined by CSF $A\beta_{42}/A\beta_{40}$ ratio only* | | | |
|---|---|---|---|
| **Lumipulse** | **Test low** | **Test indeterminate** | **Test high** |
| True CSF negative | 74 | 19 | 5 |
| True CSF positive | 8 | 29 | 122 |
| **ALZpath** | **Test low** | **Test indeterminate** | **Test high** |
| True CSF negative | 54 | 39 | 5 |
| True CSF positive | 8 | 21 | 130 |
| ***b)*** *CSF cohort: AD status determined by CSF $A\beta_{42}/A\beta_{40}$ ratio and p-tau181* | | | |
| **Lumipulse** | **Test low** | **Test indeterminate** | **Test high** |
| True CSF negative | 97 | 24 | 6 |
| True CSF positive | 7 | 40 | 83 |
| **ALZpath** | **Test low** | **Test indeterminate** | **Test high** |
| True CSF negative | 74 | 47 | 6 |
| True CSF positive | 7 | 47 | 76 |
| ***c)*** *CSF cohort: AD status determined by clinical diagnosis (informed by CSF)* | | | |
| **Lumipulse** | **Test low** | **Test indeterminate** | **Test high** |
| True CSF negative | 97 | 19 | 6 |
| True CSF positive | 7 | 42 | 89 |
| **ALZpath** | **Test low** | **Test indeterminate** | **Test high** |
| True CSF negative | 74 | 42 | 6 |
| True CSF positive | 7 | 39 | 92 |
| ***d)*** *Amyloid PET cohort: AD status determined by amyloid PET visual read* | | | |
| **Lumipulse** | **Test low** | **Test indeterminate** | **Test high** |
| True amyloid PET negative | 16 | 11 | 1 |
| True amyloid PET positive | 2 | 45 | 1 |
| **ALZpath** | **Test low** | **Test indeterminate** | **Test high** |
| True amyloid PET negative | 19 | 8 | 1 |
| True amyloid PET positive | 2 | 25 | 21 |
| ***e)*** *Amyloid PET cohort: AD status determined by amyloid PET visual read (with amyloid PET staging)* | | | |
| **Lumipulse** | **Test low** | **Test indeterminate** | **Test high** |
| Amyloid load none | 16 | 11 | 1 |
| Amyloid load mild | 1 | 8 | 0 |
| Amyloid load significant | 1 | 37 | 1 |
| **ALZpath** | **Test low** | **Test indeterminate** | **Test high** |
| Amyloid load none | 19 | 8 | 1 |
| Amyloid load mild | 2 | 5 | 2 |
| Amyloid load significant | 0 | 20 | 19 |
| ***f)*** *CSF $A\beta_{42}/A\beta_{40}$ ratio only cut-points applied to amyloid PET cohort* | | | |
| **Lumipulse** | **Test low** | **Test indeterminate** | **Test high** |
| True amyloid PET negative | 16 | 9 | 3 |
| True amyloid PET positive | 2 | 16 | 30 |
| **ALZpath** | **Test low** | **Test indeterminate** | **Test high** |
| True amyloid PET negative | 6 | 16 | 6 |
| True amyloid PET positive | 1 | 4 | 43 |
| ***g)*** *CSF $A\beta_{42}/A\beta_{40}$ ratio only cut-points applied to amyloid PET cohort (with amyloid PET staging)* | | | |

| **Lumipulse** | **Test low** | **Test indeterminate** | **Test high** |
|---|---|---|---|
| Amyloid load none | 16 | 9 | 3 |
| Amyloid load mild | 1 | 6 | 2 |
| Amyloid load significant | 1 | 10 | 28 |
| **ALZpath** | **Test low** | **Test indeterminate** | **Test high** |
| Amyloid load none | 6 | 16 | 6 |
| Amyloid load mild | 1 | 3 | 5 |
| Amyloid load significant | 0 | 1 | 38 |
| **h)** *CSF $A\beta_{42}/A\beta_{40}$ ratio and p-tau181 cut-points applied to amyloid PET cohort* | | | |
| **Lumipulse** | **Test low** | **Test indeterminate** | **Test high** |
| True amyloid PET negative | 22 | 4 | 2 |
| True amyloid PET positive | 8 | 24 | 16 |
| **ALZpath** | **Test low** | **Test indeterminate** | **Test high** |
| True amyloid PET negative | 13 | 11 | 4 |
| True amyloid PET positive | 1 | 18 | 29 |
| ***i)*** *CSF $A\beta_{42}/A\beta_{40}$ ratio and p-tau181 cut-points applied to amyloid PET cohort (with amyloid PET staging)* | | | |
| **Lumipulse** | **Test low** | **Test indeterminate** | **Test high** |
| Amyloid load none | 22 | 4 | 2 |
| Amyloid load mild | 3 | 4 | 2 |
| Amyloid load significant | 5 | 20 | 14 |
| **ALZpath** | **Test low** | **Test indeterminate** | **Test high** |
| Amyloid load none | 13 | 2 | 4 |
| Amyloid load mild | 1 | 6 | 2 |
| Amyloid load significant | 0 | 12 | 27 |
| **j)** *Clinical diagnosis (informed by CSF) cut-points applied to amyloid PET cohort* | | | |
| **Lumipulse** | **Test low** | **Test indeterminate** | **Test high** |
| True amyloid PET negative | 22 | 4 | 2 |
| True amyloid PET positive | 8 | 23 | 17 |
| **ALZpath** | **Test low** | **Test indeterminate** | **Test high** |
| True amyloid PET negative | 13 | 11 | 4 |
| True amyloid PET positive | 1 | 17 | 30 |
| ***k)*** *Clinical diagnosis (informed by CSF) cut-points applied to amyloid PET cohort (with amyloid PET staging)* | | | |
| **Lumipulse** | **Test low** | **Test indeterminate** | **Test high** |
| Amyloid load none | 22 | 4 | 2 |
| Amyloid load mild | 3 | 4 | 2 |
| Amyloid load significant | 5 | 19 | 15 |
| **ALZpath** | **Test low** | **Test indeterminate** | **Test high** |
| Amyloid load none | 13 | 11 | 4 |
| Amyloid load mild | 1 | 6 | 2 |
| Amyloid load significant | 0 | 11 | 28 |

*Abbreviations: amyloid-beta 42 to amyloid-beta 40 ratio ($A\beta_{42}/A\beta_{40}$), Alzheimer's disease (AD).*

**Supplementary Table 8:** Lumipulse and ALZpath p-tau217 cut-points and intermediate zone percentages determined by varied methods of AD status classification using varied sensitivity and specificity in the amyloid PET cohort and with CSF cohort cut-points applied to amyloid PET cohort.

| Assay | 90% Sensitivity cut-point | 90% Specificity cut-point | % in Intermediate zone | 95% Sensitivity cut-point | 95% Specificity cut-point | % Intermediate zone | 97.5% sensitivity cut-point | 97.5% specificity cut-point | % Intermediate zone |
|---|---|---|---|---|---|---|---|---|---|
| | *AD status defined by Amyloid PET visual read (amyloid PET cohort-derived cut-points)* | | | | | | | | |
| **Lumipulse** | 0.211 | 0.313 | 14.5 | 0.160 | 1.372 | 73.7 | 0.138 | 1.372 | 77.6 |
| **ALZpath** | 0.489 | 0.955 | 26.3 | 0.375 | 1.091 | 43.4 | 0.217 | 1.091 | 61.8 |
| | *AD status defined by CSF $A\beta_{42}/A\beta_{40}$ ratio only (CSF cohort-derived cut-points applied to amyloid PET data)* | | | | | | | | |
| **Lumipulse** | 0.236 | 0.254 | 1.3 | 0.150 | 0.380 | 32.9 | 0.109 | 0.664 | 63.2 |
| **ALZpath** | 0.325 | 0.419 | 7.9 | 0.220 | 0.486 | 26.3 | 0.190 | 0.558 | 34.2 |
| | *AD status defined by CSF $A\beta_{42}/A\beta_{40}$ ratio and p-tau181 (CSF cohort-derived cut-points applied to amyloid PET data)* | | | | | | | | |
| **Lumipulse** | 0.301 | 0.438 | 11.8 | 0.239 | 0.700 | 36.8 | 0.157 | 0.844 | 60.5 |
| **ALZpath** | 0.458 | 0.610 | 10.5 | 0.278 | 0.870 | 38.2 | 0.212 | 1.093 | 61.8 |
| | *AD status defined by most recent clinical diagnosis (CSF cohort-derived cut-points applied to amyloid PET data)* | | | | | | | | |
| **Lumipulse** | 0.301 | 0.403 | 10.5 | 0.236 | 0.660 | 35.5 | 0.178 | 0.763 | 54.0 |
| **ALZpath** | 0.458 | 0.558 | 6.6 | 0.278 | 0.798 | 36.8 | 0.220 | 1.042 | 59.2 |

*Abbreviations: amyloid-beta 42 to amyloid-beta 40 ratio ($A\beta_{42}/A\beta_{40}$), Alzheimer's disease (AD).*

**Supplementary Table 9:** Multiple linear regression coefficients (with 95% CI and significance levels) for age, sex, serum creatinine, CKD stage (ordinal and binary for stage 3 and above) and BMI on log-transformed Lumipulse p-tau217 measurements in the CKD and CSF cohorts combined. Data presented is from a subset of the cohort data in which all variables were available (n=80).

| Model | | | Coefficient (95% CI; p-value) | | | | |
|---|---|---|---|---|---|---|---|
| | **Cohort** | **Age** | **Sex** | **CKD stage** | **BMI** | **SCr** | **AIC** |
| **Model 1** | -0.264 (-0.850, 0.322) p=0.372 | 0.012 (-0.009, 0.033) p=0.267 | -0.327 (-0.719, -0.064) p=0.100 | - | - | - | 209.3 |
| **Model 2** (for all CKD stages) | -0.586 (-1.306, 0.134) p=0.109 | 0.007 (-0.016, 0.030) p=0.553 | -0.286 (-0.691, 0.119) p=0.163 | CKD-2: 0.237 (-0.249, 0.723; p=0.334)<br>CKD-3a: 0.537 (-0.181, 1.256; p=0.140)<br>CKD-3b: 0.271 (-0.508, 1.049; p=0.490)<br>CKD-4: 0.600 (-0.243, 1.444; p=0.160) | - | - | 213.8 |
| **Model 3** (for all CKD stages) | -0.499 (-1.207, 0.210) p = 0.165) | 0.012 (-0.012, 0.035) p=0.326 | -0.355 (-0.733, 0.064) p=0.099 | CKD-2: 0.104 (-0.388, 0.595; p=0.675)<br>CKD-3a: 0.444 (-0.264, 1.152; p=0.215)<br>CKD-3b: 0.322 (-0.441, 1.084; p=0.403)<br>CKD-4: 0.515 (-0.314, 1.343; p=0.220) | -0.039 (-0.077, -0.002) p=0.040 * | - | 211.1 |
| **Model 4** (CKD stage as binary for stage 3 and above) | -0.489 (-1.162, 0.183) p=0.152) | 0.009 (-0.013, 0.030) p=0.428 | -0.326 (-0.715, 0.064) p=0.100 | 0.331 (-0.162, 0.824; p=0.185) | - | - | 209.5 |
| **Model 5** (CKD stage as binary for stage 3 and above) | -0.459 (-1.112, 0.193) p=0.165) | 0.012 (-0.009, 0.033) p=0.255 | -0.354 (-0.733, 0.024) p=0.066 | 0.365 (-0.113, 0.844; p=0.133) | -0.042 (-0.078, -0.007) p=0.019 * | - | 205.4 |
| **Model 6** | -0.369 (-0.971, 0.234; 0.227) | 0.017 (-0.004, 0.038) p=0.111 | -0.451 (-0.850, -0.052) p=0.027) * | - | -0.040 (-0.075, -0.005) p=0.025) * | 0.003 (-0.001, 0.008; 0.133) | 205.4 |

*Note: significance level * p < 0.05. Abbreviations: Akaike information criterion (AIC), body mass index (BMI), chronic kidney disease (CKD), confidence interval (CI), serum creatinine (SCr).*

**Supplementary Table 10:** Pre-analytical handling factors statistical testing.

| | **Lumipulse** | | **ALZpath** | |
|---|---|---|---|---|
| **Experiment** | $\chi 2$ | *p* | $\chi 2$ | *p* |
| Pre-centrifugation delay | 1.385 | 0.500 | 9.800 | 0.008 ** |
| Pre-centrifugation delay at 2-8°C | 2.400 | 0.301 | 4.200 | 0.123 |
| Post-centrifugation delay | 18.711 | 0.005 *** | 7.800 | 0.253 |
| Number of freeze-thaw cycles | 1.485 | 0.686 | 3.000 | 0.392 |

*After statistically significant results on Friedman tests, pairwise post-hoc Nemenyi testing was conducted to identify the source of the between-group difference (** $p < 0.01$, *** $p < 0.005$).*
